# Mental health in the UK Biobank: a roadmap to self-report measures and neuroimaging correlates

**DOI:** 10.1101/2021.04.08.21255070

**Authors:** Rosie Dutt, Kayla Hannon, Ty Easley, Joseph Griffis, Wei Zhang, Janine Bijsterbosch

**Author notes:** Corresponding author: Janine Bijsterbosch. Joint first authors (Rosie Dutt, Kayla Hannon).

## Abstract

The UK Biobank (UKB) is a highly promising dataset for brain biomarker research into population mental health due to its unprecedented sample size and extensive phenotypic, imaging, and biological measurements. In this study, we aimed to provide a shared foundation for UKB neuroimaging research into mental health with a focus on anxiety and depression. We compared UKB self-report measures and revealed important timing effects between scan acquisition and separate online acquisition of some mental health measures. To overcome these timing effects, we introduced and validated the Recent Depressive Symptoms (RDS-4) score which we recommend for state-dependent and longitudinal research in the UKB. We furthermore tested univariate and multivariate associations between brain imaging derived phenotypes (IDPs) and mental health. Our results showed a significant multivariate relationship between IDPs and mental health, which was replicable. Conversely, effect sizes for individual IDPs were small. Test-retest reliability of IDPs was stronger for measures of brain structure than for measures of brain function. Taken together, these results provide benchmarks and guidelines for future UKB research into brain biomarkers of mental health.

## 1. Introduction

Over the years there have been a multitude of neuroimaging studies that aimed to investigate alterations in the brain in relation to affect-based mental health (e.g., anxiety and depression). The Major Depressive Disorder (MDD) literature reports structural changes in the cortico-limbic network [Klauser et al., 2015], insula and hippocampus [Stratmann et al., 2014], as well as functional changes in the Default Mode Network (DMN) [Tozzi et al., 2021; Yu et al., 2019], medial temporal gyrus, and caudate [Ma et al., 2012]. In Generalized Anxiety Disorder (GAD), similar functional changes are seen in the DMN [Andreescu et al., 2011] and ventromedial prefrontal cortex [Cha et al., 2014], as well as structural changes in the DMN [Wolf et al., 2016] and amygdala [He et al., 2016]. However, the literature on neural correlates of MDD contains some inconsistent findings. For example, some studies report greater functional connectivity in the DMN [Greicius et al., 2007; Sheline et al., 2010] while others report lesser functional connectivity in the same network [Bluhm et al., 2009; Tozzi et al., 2021; Yan et al., 2019]. A potential reason for inconsistent findings is the small sample size of most of these studies. The broader fields of psychology and neuroimaging are recognizing that small sample sizes lead to inflated effect sizes that often result from sampling variability and therefore do not replicate in new data [Button et al., 2013; Grady et al., 2021; Marek et al., 2020; Poldrack et al., 2017; Yarkoni, 2009]. Larger sample sizes are therefore needed to obtain reliable insights into the neural correlates of mental health.

One option to achieve larger sample sizes is to conduct meta-analyses. Meta-analyses use results from prior studies as their input and employ quantitative methods to pool data across studies and test for consensus [Müller et al., 2018]. A meta-analysis on resting-state functional connectivity in MDD showed hypo-connectivity in fronto-parietal and salience networks and hyper-connectivity in the DMN [Kaiser et al., 2015]. Another meta-analysis showed that there are common grey-matter volume changes in MDD which are also seen in bipolar disorder [Wise et al., 2017b]. In GAD, meta-analyses have also been able to confirm consistent dysregulation of affective control related to numerous networks, which provides support for an integrated model of brain network changes [Xu et al., 2019]. Whilst these meta-analyses aid to establish consensus on brain correlates of mental health [Wager et al., 2007], they can be limited in their scope. This is because the input studies surveyed in meta analyses often adopt narrow inclusion and exclusion criteria for the patient sample, which limits cross-diagnostic mental health research. Additionally, due to the lack of availability of whole brain statistical result images from prior studies, coordinate-based meta-analyses are often undertaken which are limited in their spatial precision [Müller et al., 2018]. Furthermore, meta analyses suffer from publication bias (only including effect sizes from published significant studies) [Thornton and Lee, 2000], language bias (only including papers written in English) [Egger et al., 1997], and selective outcome reporting (input-papers selectively publish only significant variables) [Hutton and Williamson, 2000; Kirkham et al., 2010], which can lead to inflated meta analytical results [Sterne et al., 2001]. These inherent limitations of meta-analyses may explain why disagreement persists within even meta-analytical work, with a recent study showing hypo-(rather than hyper-) connectivity in the core DMN in patients with depression [Tozzi et al., 2021].

Consequently, in recent years there has been a move to accrue larger neuroimaging datasets such as the Young Adult and Lifespan Human Connectome Projects (HCP) [Harms et al., 2018; Van Essen et al., 2013], Connectomes Related to Human Disease studies (CRHD) [Tozzi et al., 2020], UK Biobank (UKB) [Miller et al., 2016; Sudlow et al., 2015], Enhancing Neuro Imaging Genetics through Meta-Analysis ENIGMA) [Schmaal et al., 2017], and Adolescent Brain Cognitive Development study (ABCD) [Casey et al., 2018]. The increased statistical power afforded by these datasets enables studies to approximate the true effect [Marek et al., 2020]. Currently, the UKB is the largest neuroimaging dataset, encompassing data from extensive questionnaires, physical and cognitive measures, and biological samples (including genotyping) in addition to multimodal neuroimaging scans [Sudlow et al., 2015]. The UKB is a prospective epidemiological study that recruited a cohort of 500,000 participants, of which 100,000 subjects will take part in one round of imaging, and 10,000 of those subjects will undergo a further second round of scanning [Sudlow et al., 2015]. Health outcomes for all participants will be tracked over future years until participants’ decease, including full primary health and hospital records. Therefore, the UKB offers a valuable resource to study mental health and other disorders. The goal of our study is to establish a foundation for future mental health biomarker research in the UKB.

The UK Biobank includes multiple rich self-report measures of mental health. However, the organization and abundance of this information can make it somewhat challenging for researchers to navigate. For data pertaining to mental health, there are three sources within the UK Biobank. The first are assessment center questions (https://biobank.ndph.ox.ac.uk/showcase/label.cgi?id=100060) which participants complete via a touch screen on the day they were scanned. The second is a separately administered online mental health questionnaire (https://biobank.ndph.ox.ac.uk/showcase/label.cgi?id=136), which is completed by a subset of UKB participants at a time independent from the scanning date (median absolute number of days between scan 1 and online questionnaire completion: 742, range: -1,185 to +964 days in exploratory sample). The third are the health records available in the UKB which encompass the date of the first experience of specific ICD-10 diagnoses obtained from primary care (https://biobank.ndph.ox.ac.uk/showcase/label.cgi?id=3000) and hospital inpatient data (https://biobank.ndph.ox.ac.uk/showcase/label.cgi?id=2000). In this study, we tabulate and compare different mental health measures available in the UKB, with a focus on self-reported symptom scores from the assessment center information and online questionnaire. We test their relationship with brain measures, thereby providing a benchmark for using UKB mental health variables in future research.

This study aims to achieve four key goals. Firstly, we aim to clearly tabulate the different self-report measures of mental health available in the UKB and discern the relationships between summary scores to enable future studies to make an informed decision on which measure is most appropriate to use. Secondly, we propose and validate a new summary measure (Recent Depressive Symptoms; RDS-4) that uses depression questions which were asked on the day of scanning in the UK Biobank study. The RDS-4 score therefore enables research into current depressive symptoms and changes in symptomatology over time. Thirdly, we aim to establish realistic and robust univariate and multivariate effect sizes of commonly reported brain correlates of mental health based on population data. Lastly, we aim to determine the test-retest reliability of imaging variables alongside their effect size as both reliability and sensitivity are critical requirements for biomarker research. Large-scale imaging datasets such as the UKB play a critical role in the long-term goal of finding brain biomarkers of mental health, and our hope is to provide a foundation that future studies can build on.

## 2. Methods

### Dataset

Imaging data from 32,420 UKB participants were available at the time the study was performed. From this we selected multiple independent test cohorts (Fig. 1; Table 1). Subjects with a mean head motion greater than 0.2mm were removed resulting in the exclusion of 5,265 subjects. Subjects with any missing online questionnaire or scan 1 assessment center mental health data were also removed, resulting in the exclusion of additional 10,848 subjects (largely because the online questionnaire was only performed in a subset of UKB participants). From the remaining 16,307 subjects, we selected individuals who had undergone brain scans at two timepoints. These subjects make up the test-retest sample.

**Table 1.** Demographics for samples. SD = Standard Deviation. The ‘ever seen GP for mental health’ and ‘never seen GP for mental health’ subjects were matched, such that the same male-to-female ratio and mean age applies to these groups.

| Sample | N | Sex (n male) | Age (mean±SD) | Time between scans (mean absolute days±SD) |
| --- | --- | --- | --- | --- |
| Exploratory | 6,636 | 2,258 | 61.9±7.2 | N.A. |
| Confirmatory | 2,426 | 796 | 60.6±7.1 | N.A. |
| Test-Retest | 624 | 300 | 61.7±7.04 | 823.7±44.8 |

**Figure 1:**
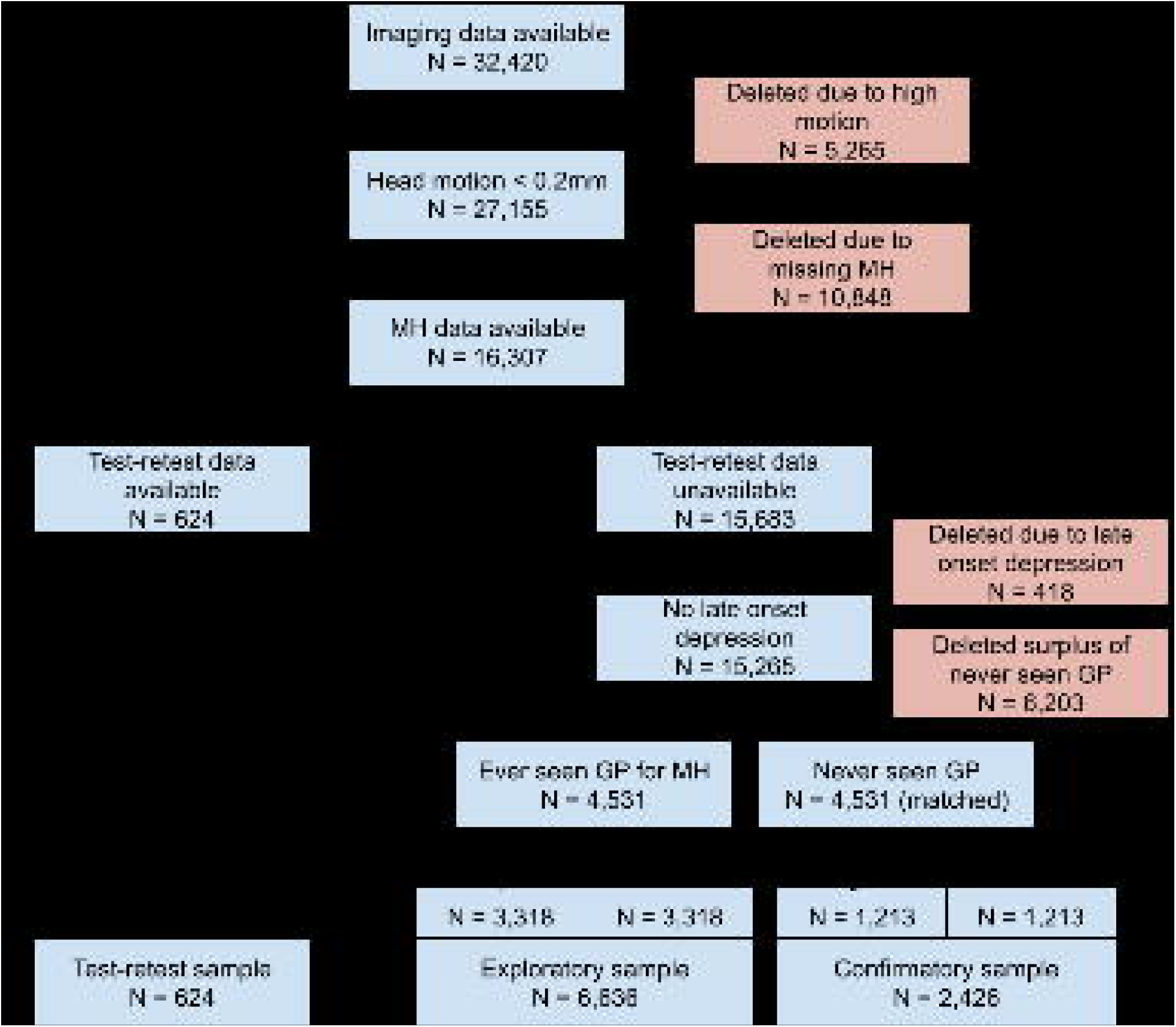
UK Biobank subject inclusion chart.

Late onset depression (first episode at age 60+) is associated with different brain correlates (e.g., white matter hyperintensities) and different risk factors (e.g., vascular risk) compared with recurrent early onset depression (age of first episode before 60) [Salo et al., 2019]. Therefore, we assessed subjects for probable late-onset depression based on self-reported age at the first episode of depression (Data-field: 20433). Subjects who reported their first episode at 60 or older (N=418) were excluded.

The majority of individuals within the UKB cohort are expected to have no mental health conditions because it is a population sample. To ensure sufficient power to identify neural correlates of mental health, we wanted to reduce the expected over-representation of healthy individuals and ensure that our samples richly capture mental health variability. This was achieved by including equal numbers of participants with and without a history of mental health. From the UKB showcase we used: Seen doctor (GP) for nerves, anxiety, tension or depression (Data-field: 2090) to ensure our samples included an equal number of subjects who experienced mental health issues on at least one occasion, and those who have not. For each subject who had seen a GP for nerves, anxiety, tension or depression (N=4,531) we paired a matched subject from those who had never seen a GP for nerves, anxiety, tension or depression (i.e., subject pairs were identically matched for sex and age, and minimal difference in head motion). Subsequently, approximately two-thirds of the ‘never seen GP’ subjects together with their matched ‘seen GP’ subjects was randomly assigned to the exploratory sample, and the remaining subjects were assigned to the confirmatory sample. During subject assignment to groups, we preserved the matched characteristics within each resulting sample (Fig. 1). No subjects overlapped between the exploratory and confirmatory samples.

### Mental health measures

The set of self-report questions related to mental health included in the UKB were informed by standardized measures, but did not simply cover a list of previously validated scales. Table 2 summarizes the 5 different UKB mental health measures, which will be used for neuroimaging and questionnaire comparison analyses and Fig. 2 provides an overview of the acquisition timing of these mental health measures relative to the scan days. The questions included in the online questionnaire enable calculation of the **Generalized Anxiety Disorder (GAD-7)** and **Patient Health Questionnaire (PHQ-9)** scores [Davis et al., 2020]. Using the Assessment center information, the Eysenck **Neuroticism (N-12)** score was calculated. Smith and colleagues used questions from the Assessment information to develop a categorical (case-control) measure of depression [Smith et al., 2013]. For the purposes of our study we adopted similar definitions to obtain a categorical assignment of **Probable Depression Status**, but we did not differentiate between single and recurrent episodes of depression. Depression status was set to 1 if subjects responded yes to variable IDs 4598 or 4631 (ever depressed | ever unenthusiastic/disinterested), *and* reported a duration of at least 1 week to variable IDs 4609 or 5375 (depression | unenthusiasm/disinterest), *and* had seen either a GP or psychiatrist for nerves, anxiety, tension, depression (i.e., responded yes to variable IDs 2090 or 2100).

**Table 2:** Measures of affect-based mental health available in the UK Biobank. PHQ-9 = Patient Health Questionnaire-9, RDS-4 = Recent Depressive Symptoms-4, GAD-7 = General Anxiety Disorder-7, N-12 = Neuroticism-12.

|  | Scan day | Online | Range | Questions | Variable IDs |
| --- | --- | --- | --- | --- | --- |
| PHQ-9 |  | ✓ | 0-27 | Little interest or pleasure in doing things<br>Feeling down, depressed, or hopeless<br>Trouble sleeping<br>Feeling tired<br>Poor appetite or overeating<br>Feeling bad about yourself<br>Trouble concentrating<br>Moving or speaking slowly or fidgety or restless | 20514<br>20510<br>20517<br>20519<br>20511<br>20507<br>20508<br>20518 |
|  |  |  |  | Thoughts that you would be better off dead | 20513 |
| RDS-4 | ✓ |  | 4-16 | Frequency of depressed mood in last 2 weeks | 2050 |
|  |  |  |  | Frequency of unenthusiasm / disinterest in last 2 weeks | 2060 |
|  |  |  |  | Frequency of tenseness / restlessness in last 2 weeks | 2070 |
|  |  |  |  | Frequency of tiredness / lethargy in last 2 weeks | 2080 |
| GAD-7 |  | ✓ | 0-21 | Feeling nervous, anxious or on edge | 20506 |
|  |  |  |  | Not being able to stop or control worrying | 20509 |
|  |  |  |  | Worrying too much about different things | 20520 |
|  |  |  |  | Trouble relaxing | 20515 |
|  |  |  |  | Being so restless that it is hard to sit still | 20516 |
|  |  |  |  | Becoming easily annoyed or irritable | 20505 |
|  |  |  |  | Feeling afraid as if something awful might happen | 20512 |
| N-12 | ✓ |  | 0-12 | Mood swings | 1920 |
|  |  |  |  | Miserableness | 1930 |
|  |  |  |  | Irritability | 1940 |
|  |  |  |  | Sensitivity / hurt feelings | 1950 |
|  |  |  |  | Fed-up feelings | 1960 |
|  |  |  |  | Nervous feelings | 1970 |
|  |  |  |  | Worrier / anxious feelings | 1980 |
|  |  |  |  | Tense / 'highly strung' | 1990 |
|  |  |  |  | Worry too long after embarrassment | 2000 |
|  |  |  |  | Suffer from 'nerves' | 2010 |
|  |  |  |  | Loneliness, isolation | 2020 |
|  |  |  |  | Guilty feelings | 2030 |
| Probable depression status | ✓ |  | 0/1 | Ever depressed | 4598 |
|  |  |  |  | Ever unenthusiastic/disinterested | 4631 |
|  |  |  |  | Duration of longest period of depression | 4609 |
|  |  |  |  | Duration of longest period of unenthusiasm/disinterest | 5375 |
|  |  |  |  | Seen Doctor (GP) for nerves, anxiety, tension, depression | 2090 |
|  |  |  |  | Seen psychiatrist for nerves, anxiety, tension, depression | 2100 |

**Figure 2:**
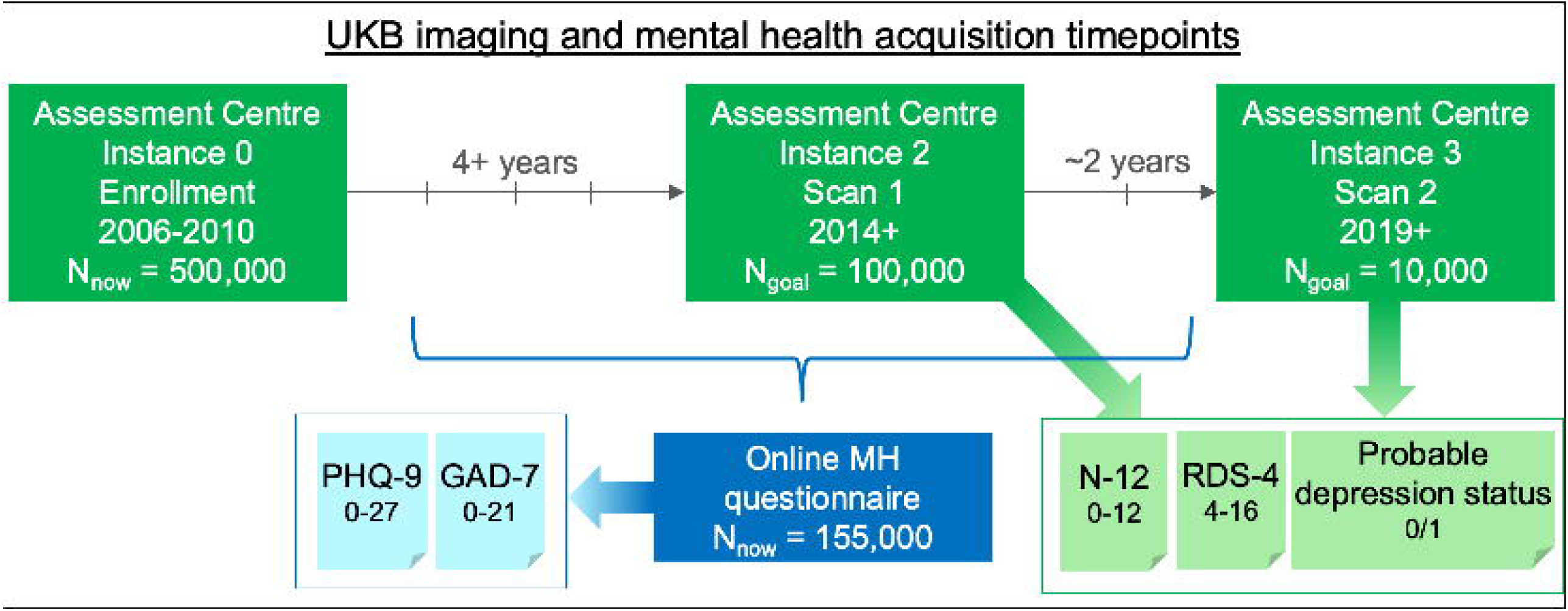
Schematic overview of the acquisition timing of UK Biobank mental health measures in relation to imaging acquisition. Mental health measures in light green were obtained on the day of scanning, whereas mental health measures in light blue were obtained at an independent time point that varied from 1,185 days before to 964 days after scan 1 across participants. The range of possible scores for each mental health measure is included. All five measures were included in neuroimaging and questionnaire comparison analyses in this paper.

For our study, we proposed a new summary measure of state depression using UKB questions included in the Assessment center information: **Recent Depressive Symptoms (RDS-4)**, which is a continuous measure of depression symptomatology obtained on the day of scanning. The four self-report questions used for the RDS-4 assess depressed mood, disinterest, restlessness, and tiredness. Each question asks about recent experiences of symptoms (past 2 weeks). The response options for the four questions are: not at all (1), several days (2), more than half the days (3), and nearly every day (4). The summed score across these four variables therefore has a range of 4-16. Moreover, the RDS-4 questions correspond with several DSM-V diagnostic criteria for major depressive disorder and cover depression domains that are also considered in other measures such as the Hamilton and Montgomery–Åsberg scales.

There are a number of important differences between the RDS-4 and the other mental health measures. Compared to PHQ-9, the RDS-4 was obtained on the day of the imaging scan, whereas the PHQ-9 was undertaken at a time point that was independent from the scan date. Compared to probable depression status, the RDS-4 provides a continuous measure of recent symptom severity, whereas probable depression status is a categorical (case-control) measure of lifetime occurrence of depression. Compared to N-12, RDS-4 is a measure of recent (‘state’) depressive symptoms, whereas N-12 is a more general measure of personality (‘trait’). Compared to GAD-7, the RDS-4 focuses on depression and the GAD-7 focuses on anxiety.

### Imaging acquisition

UKB structural modalities include: T1-weighted (T1), T2-weighted (T2), susceptibility-weighted MRI (swMRI); diffusion MRI (dMRI); and functional modalities: task-based fMRI (tfMRI) and resting-state fMRI (rsfMRI). MRI data were obtained using a Siemens Magnetom Skyra 3T scanner. For T1 structural scans, 3D MPRAGE acquisition was used to acquire 1mm isotropic resolution. For T2 scans fluid-attenuated inversion recovery (FLAIR) contrast was used with the 3D SPACE optimized readout providing a strong contrast for white matter hyperintensities. For swMRI, a 3D gradient echo acquisition was used (resolution: 0.8×0.8×3mm), obtaining two echo times (TE=9.4 and TE=20 ms). Diffusion data was acquired with b-values of 1000 and 2000 s/mm2, at 2mm spatial resolution, with a factor 3 multiband acceleration and 50 distinct diffusion-encoding directions. Both tfMRI and rs-fMRI used identical acquisition parameters (spatial resolution= 2.4mm, TR= 0.735s, factor = 8 multiband accelerator). Task fMRI used the Hariri faces/shapes “emotion” task as employed in the HCP [Barch et al., 2013; Hariri et al., 2002], with a shorter total length and reduced repeats of the total stimulus block. For further information on UKB imaging, please refer to [Miller et al., 2016].

### Imaging derived phenotypes

In addition to raw and processed imaging data, Image Derived Phenotypes (IDPs) are available for download. IDPs are derived from calculations that combine many images and/or voxels to produce a scalar quantity from the processed imaging data [Miller et al., 2016]. Examples of IDPs include regional volumes from structural MRI and ‘edges’ from resting state functional MRI (i.e., connectivity between a pair of networks).

The IDPs included in this paper are summarized in Table 3, and further information can be found in [Miller et al., 2016] as well as the UKB showcase brain imaging documentation resource (https://biobank.ndph.ox.ac.uk/showcase/showcase/docs/brain_mri.pdf). Briefly, resting state IDPs were obtained using Independent Components Analysis performed at two different dimensionalities (25 and 100), which resulted in 21 and 55 signal networks, respectively. Subject-specific BOLD time series for each network were calculated using dual regression [Nickerson et al., 2017], and the amplitude for each network (temporal standard deviation) and functional connectivity between pairs of networks (full or partial correlation coefficients) were calculated. Resting state IDPs from both ICA dimensionalities were included as they may offer complimentary information at different levels of functional organization. From T1-weighted images, gray matter volumes were obtained with FSL FIRST and FAST, and cortical area and thickness were calculated with Freesurfer. Total volume of white matter hyperintensities was estimated based on T1-weighted and T2-flair images using FSL’s BIANCA algorithm [Griffanti et al., 2016]. From the diffusion data, weighted mean fractional anisotropy (FA) and mean diffusivity (MD) were obtained using FSL’s DTIFIT tool. Task fMRI IDPs reflect summary measures of activation (the median and 90th percentile for both the percent signal change and the z-statistic) in regions selected from the group-level activation map. Susceptibility weighted IDPs were generated from the signal decay times predicted from the magnitude images at the two TEs such that the IDPs equate to the median signal decay times.

**Table 3:** Full set of IDPs considered for canonical correlation analysis. IDP = Imaging Derived Phenotype, UKB = UK Biobank.

|  | # IDPs | UKB ID | Description |
| --- | --- | --- | --- |
| **Resting state** | 21 | 25754 | rfMRI network amplitudes from 21 signal components |
|  | 55 | 25755 | rfMRI network amplitudes from 55 signal components |
|  | 210 | 25750 | Pairwise full correlation edges between 21 components |
|  | 210 | 25752 | Pairwise partial correlation edges between 21 components |
| **Total 3,466** | 1,485 | 25751 | Pairwise full correlation edges between 55 components |
|  | 1,485 | 25753 | Pairwise partial correlation edges between 55 components |
| **Structural** | 139 | 1101 | FAST gray matter volumes |
|  | 14 | 1102 | FIRST gray matter volumes |
|  | 62 | 196 | Cortical surface area from Freesurfer DKT atlas |
|  | 62 | 196 | Cortical thickness from Freesurfer DKT atlas |
|  | 1 | 25781 | Total volume of white matter hyperintensity |
|  | 27 | 107 | Weighted-mean FA |
| **Total** | 27 | 107 | Weighted-mean MD |
| **346** | 14 | 109 | Median T2-star from susceptibility weighted imaging |
| **Task** | 16 | 106 | Task fMRI median + 90th percentile of BOLD effect and z |

### Confound variables

All analyses were corrected for the ‘simple’ set of confounds described in [Alfaro-Almagro et al., 2021], namely: scanning site, age, age squared, sex, age * sex, head size, head motion in resting fMRI and in task fMRI scans, date, and date squared. This confound set was previously shown to explain 4.4% of variance in UKB imaging variables on average, and captured the most important sources of confound variation [Alfaro-Almagro et al., 2021].

### Correlations amongst mental health variables in the UKB

To characterize the degree of overlapping information between mental health measures, Spearman rank correlations were computed between all measures of mental health using data from the exploratory sample (N=6,636).

Data used to compute RDS-4 and N-12 were collected at the scan date (assessment center information), whereas GAD-7 and PHQ-9 were computed from data obtained from the online questionnaire. The absolute number of days elapsed between the two data collections ranged from 0 to 1,185 days. To investigate the effects of measurement latency on mental health measure correlation, Spearman rank correlations between the RDS-4 and PHQ-9 (both measures of depression) were computed as a function of elapsed time between measurement (see Supplementary Materials section S1 and Fig. S1).

To test whether self-report measures differed significantly based on probable depression status, a two-sample Kolmogorov-Smirnov test was performed to ascertain whether subjects with a positive depression status had different distributions of depression scores than subjects with no depression status.

### Mapping between mental health variables in the UKB

To gain insights into how the different measures of mental health included in the UKB relate to each other, we used equipercentile linking in the exploratory sample. Here, the stepwise percentiles for each measure were calculated, and for each score in one measure the equivalent percentile rank in a different measure was mapped [Kolen and Brennan, 2014]. We further calculated the Cronbach alpha for the newly proposed RDS-4 score to measure internal consistency in the exploratory sample.

### Mechanical Turk study to validate RDS-4

To further validate the proposed RDS-4 score, we performed an independent study using the Amazon Mechanical Turk platform via CloudResearch.com [Litman et al., 2017]. Participants were paid a nominal compensation for questionnaire completion. 134 participants aged 60+ completed the study. This study was reviewed by the Washington University in St Louis IRB board and approved as exempt (IRB #201909165) because participants were fully anonymous (the option of ‘anonymized worker IDs in CloudResearch was adopted) and no participant key was available to any member of the research team.

Participants completed the same set of mental health questionnaires at two timepoints 7 days apart using the Qualtrics software (Qualtrics, Provo, UT). The following questionnaires were presented in randomized order: RDS-4, PHQ-9, CES-D (Center for Epidemiological Studies - Depression; [Radloff, 1977]) and MASQ-30 (short-form Mood and Anxiety Symptoms Questionnaire; [Wardenaar et al., 2010; Watson and Clark, 1991]). The latter two measures were included because they are commonly used measures of depression that can be considered ‘gold standard’ for self-report. Although these measures are not available in the UKB, our goal was to validate the RDS-4 against these standardized measures.

We undertook multiple steps to avoid low quality responses, which can be a concern in Mechanical Turk questionnaire research. Firstly, we adopted premium options in CloudResearch, such as only including ‘CloudResearch approved participants’ who undergo more extensive vetting. Secondly, we included two questions to assess the attention levels of the participants while performing the study (*“If you are still paying attention, please select ‘yes’”* & *“Please answer this question with the ‘Most or all of the time’ option”*). Participants who failed to answer these questions appropriately were excluded. Thirdly, we imposed a minimum duration for questionnaire completion at 172.5 seconds (which equals 2.5 seconds per question). Participants who completed the questionnaire in less than 172.5 seconds were excluded.

Spearman rank correlation was used to compare scores between the RDS-4, PHQ-9, CES-D, and MASQ-30 using data from time point 1. Intraclass correlation coefficient (ICC A,1; also known as criterion-referenced reliability [Koo and Li, 2016; McGraw and Wong, 1996]) was used to calculate the test-retest reliability between time point 1 and time point 2 separately for each measure.

### Exploratory brain-mental health analysis

We used Canonical Correlation Analysis (CCA) as a data-driven approach to identify joint multivariate relationships between mental health measures and brain imaging variables [Hotelling, 1936]. Following nuisance regression to remove variance explained by nuisance regressors, dimensionality reduction was performed separately for resting state, structural, and task IDPs (Table 3) using Principal Component Analysis (PCA). The substantial differences in IDP numbers between resting state IDPs (3,466), structural IDPs (346), and task fMRI IDPs (16) was the reason for performing the dimensionality reduction separately to ensure that all classes of IDPs were represented in the input components. The top components explaining at least 50% of variance were retained for each of resting state, structural, and task IDPs. This threshold was chosen as a good trade-off between retaining a substantial amount of IDP variance for the CCA while limiting the number of input variables to the CCA to ensure a sufficient subject-to-variable ratio required for stable CCA results [Helmer et al., 2021]. The structural and task IDP matrix included a small number of missing values, which were excluded for the nuisance regression and then imputed using nearest neighbor imputation (Matlab’s knnimpute.m). The combined set of IDP eigenvectors were entered into the CCA against 5 mental health input variables corresponding to summary scores from GAD-7, N-12, PHQ-9, RDS-4 and probable depression status (residuals after regressing out confound variables). CCA was performed on N=6,636 subjects in the exploratory sample. Permutation testing with 2000 permutations was used to obtain p-values for the resulting canonical correlations. Here, the subject order of IDP component inputs and mental health inputs were independently shuffled to break subject correspondence. This is especially important for CCA because the canonical correlation is explicitly maximized and therefore it is important to compare the canonical correlation to the empirical null distribution obtained with permutation testing (which does not center around zero but shows relatively high null correlations) [Smith et al., 2015].

To calculate the univariate contributions (or ‘loadings’) from individual IDPs to the CCA result, we correlated subject scores against original IDPs. For this purpose, the ‘U’ and ‘V’ canonical subject scores from the strongest CCA result were averaged within each subject to obtain a CCA summary subject score (UV). Here, U = XA and V = YB, where X are the IDP principal component inputs and Y are the mental health inputs. A and B are the canonical coefficients for IDP eigenvectors and mental health variables respectively, which are optimized such that the correlation between U and V is maximized. We could calculate IDP contributions by correlating U with the IDPs, but the resulting correlations would potentially be inflated because U is optimized for X. Therefore, using the averaged UV subject score for correlations with the IDPs provides a more realistic and unbiased measure of individual IDP correlations [Bijsterbosch et al., 2018]. Bonferroni correction for multiple comparisons was performed for these post-hoc correlations that were used to estimate univariate contributions from each original IDP (i.e., p-value below 0.05/(3,466+346+16)=1.3*10^−5^, where 3,466 is the number of resting state IDPs, 346 is the number of structural IDPs, and 16 is the number of task IDPs). IDPs that survived correction were selected for subsequent tests of effect size in the confirmatory sample. These IDPs are referred to as ‘selected brain variables’ in subsequent confirmatory analyses.

The multivariate CCA results were also replicated in the independent confirmatory sample by projecting the resting state, structural, and task IDPs onto the same PCA subspace (i.e., not repeating the PCA, but using the weights from the exploratory sample), and multiplying brain eigenvectors as well as mental health scores by their respective canonical coefficients (i.e., A & B as estimated from the exploratory sample). The CCA replication was tested based on the correlation between the resulting U and V (i.e., the canonical correlation). We also performed the same post-hoc univariate correlations between averaged UV and individual IDPs as described above to assess the replicability of IDP contributions to the CCA.

### Confirmatory analysis of effect size

The independent confirmatory sample (N=2,426) was used to test univariate effect sizes of selected brain variables from CCA analysis (i.e., significant IDPs after Bonferroni correction). Specifically, we performed a Cohen’s d test based on probable depression status, and calculated the Pearson’s r from the correlations between the selected brain variables and each of the four mental health variables (i.e., RDS-4, PHQ-9, N-12 and GAD-7), respectively. These analyses were repeated for each imaging modality including surface area, gray matter volume, cortical thickness, white matter hyperintensity, fractional anisotropy, median T2*, task activity, resting-state network amplitude and edge connectivity at both dimensionalities (i.e., 25 and 100). We de-confounded both the brain variables and the mental health variables before running the aforementioned analyses. The only exception from de-confounding is the binary grouping based on probable depression status, as deconfounding would result in subject-specific values that are non-categorical, which is unsuitable for the Cohen’s d test.

### Test-retest reliability of imaging measures

To assess the stability of IDPs across time, we performed test-retest reliability analyses using data from N=624 subjects that were scanned twice at separate time-points, with an inter-scan interval of approximately 2 years (Table 1). Data were de-confounded for this sample using the same approach employed for the exploratory CCA analysis. After data were de-confounded, intra-class correlations were computed between the IDPs collected at each scan time-point using the ICC(A,1) formulation to quantify the agreement between measurements collected at each timepoint [McGraw and Wong, 1996]. Test-retest reliability measures were grouped according to IDP measurement modality (e.g., cortical area, cortical volume, etc.) to allow for assessment of the ICC distributions for different modalities.

We also assessed the effect of inter-scan interval length on the test-retest correlation strengths by computing ICCs for each IDP after including regressing out the inter-scan interval (in days) from each IDP, thus removing any additional variance attributable to inter-subject differences in inter-scan interval lengths. Finally, we assessed whether ICCs were affected by mental health changes as indicated by the difference in the RDS-4 scores between time-points. Of the *N*=624 subjects included in the test-retest analyses, *n*=336 exhibited no change in RDS-4 scores between time-points, while *n*=288 exhibited changes in RDS-4 scores between time-points (i.e., at least 1 point difference in the RDS-4 scores). For these analyses, we separately computed ICCs for each mental health sub-group and then plotted the ICC distributions for each modality between the sub-groups. We also computed ICCs after regressing out mental health change values from each IDP.

## 3. Results

### Correlations amongst mental health variables in the UKB

Mental health measures showed moderate correlations with one another, indicating redundancy between these metrics (Fig. 3A). RDS-4 and N-12, which are both measured from questions administered on the scan date, had a Spearman rank correlation coefficient (SRCC) of *ρ*=0.57±0.01 (*p* ≈ 10^−199^); PHQ-9 and GAD-7, which were both taken from the online questionnaire, have SRCC *ρ*=0.69±0.01 (*p* ≈ 10^−267^). Correlations between PHQ-9 and GAD-7 scores were significantly higher than between any other pairs of scores (*p*< 10^−9^).

**Figure 3.**
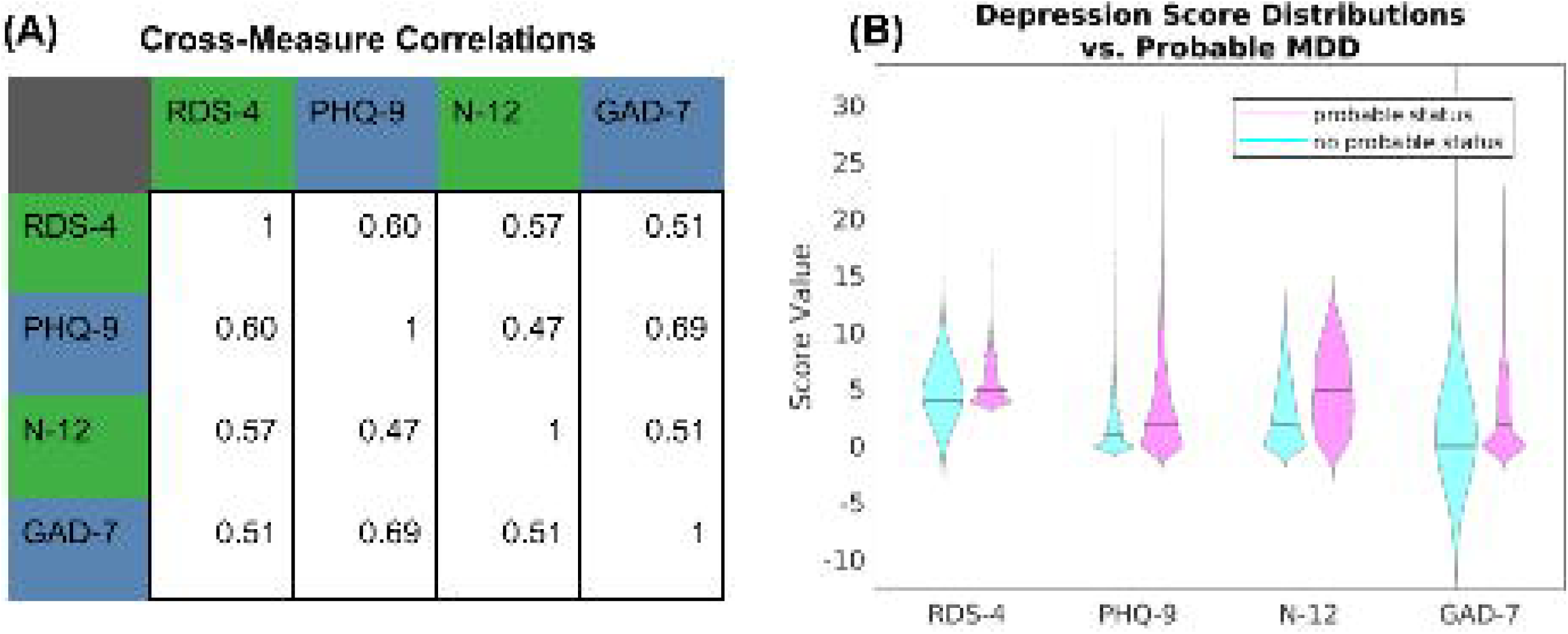
A) Spearman rank correlation coefficients between each pair of mental health measures. Variables measured on the same date are labeled the same color (green = assessment center day-of-scan information; blue = online questionnaire). B) Distributions of scores for subjects with probable depression status (pink) and without probable depression status (cyan). Subjects with probably depression status scores significantly higher on all mental health measures (KS-statistic *χ* > 0.19, *p* < 10^−48^).

Both the RDS-4 and N-12 measures were collected at each scan time, which allows for an assessment of the within-measure two-year correlation of these measures on a sample of N=555 subjects from the test-retest sample (69 subjects were removed from the full N=624 test-retest sample due to missing mental health assessment center information on scan 2). Within this subgroup, the subjects’ RDS-4 measures showed a 2-year Spearman rank correlation coefficient of *ρ* = 0.57 between initial and follow-up scans, and N-12 showed a 2-year correlation of *ρ* = 0.85. It should be noted that this reflects correlation between scan timepoints between 761 and 980 days apart. Therefore, a given metric’s 2-year correlation (i.e., self-correlation over a long time period) effectively establishes an approximate upper bound on any correlation value between it and other metrics collected over the same time frame. Because anxiety and depression are not fixed states and scores may meaningfully differ between the two timepoints available in the UKB, we also performed a separate Mechanical Turk study to test the short-term (7-day) test-retest reliability of RDS-4 (see ‘Mechanical Turk study to validate RDS-4’ section).

We performed a two-sided, two-sample Kolmogorov-Smirnov test on RDS-4, PHQ-9, N-12, and GAD-7 scores over subjects with and without probable depression status. Subjects with probable depression scored significantly higher than subjects with no probable depression status on all measures (KS-statistic *χ* > 0.19, *p* < 10^−48^; Fig. 2B).

### Mapping between mental health variables in the UKB

Given that this is a largely healthy sample, as expected, the distributions for PHQ-9, RDS-4, and GAD-7 all reveal a large number of participants with scores on the lower end of the mental health measure, with a sharp decline seen in the number of participants scoring on the upper end of the mental health measures (Fig. 4A-D). Notably, the distribution of N-12 is relatively less skewed than PHQ-9, RDS-4, and GAD-7.

**Figure 4.**
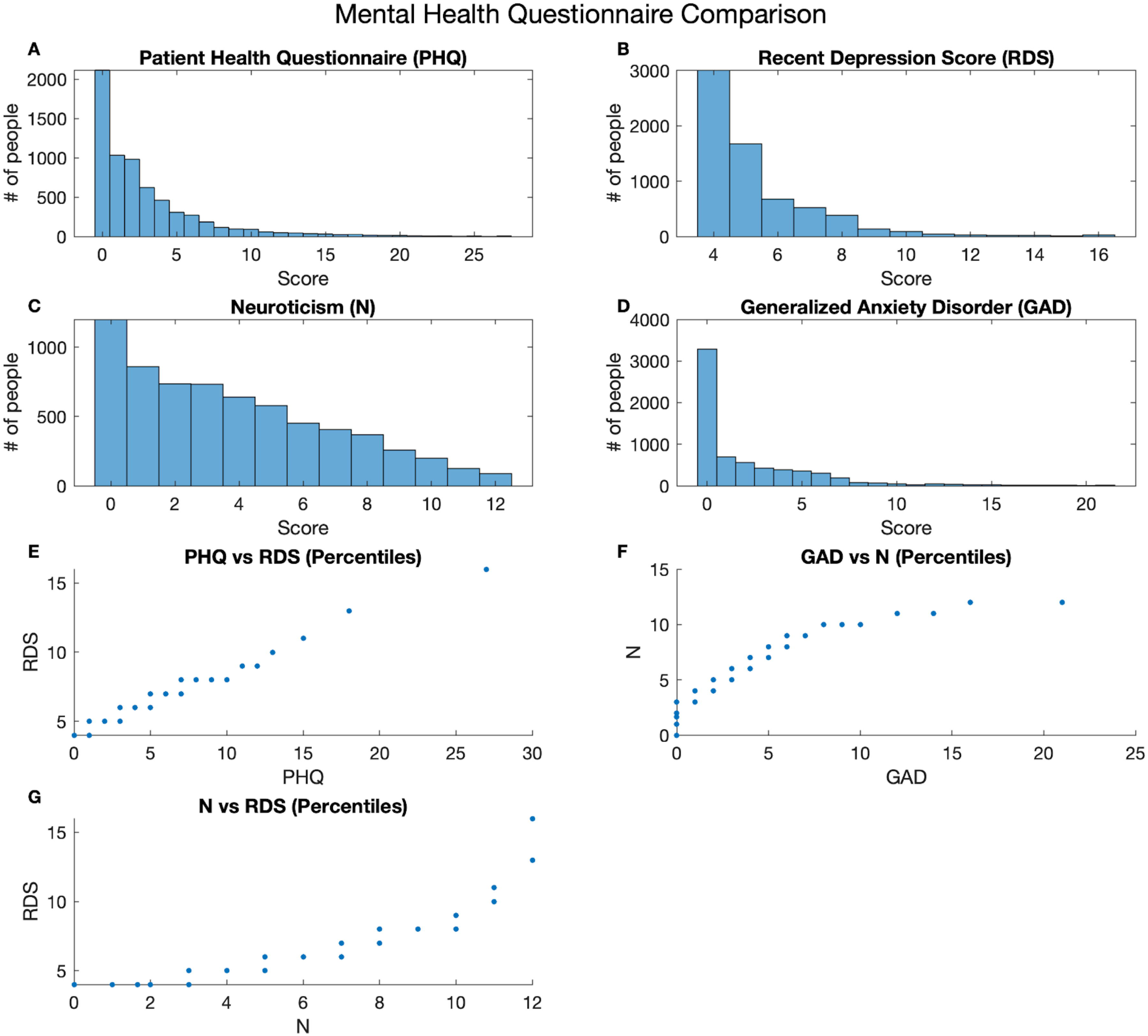
A-D are the distributions of scores for participant responses to each questionnaire. E-G depict the equipercentile linkages of the scores for each questionnaire, mapping the equivalence of a score from one questionnaire to the score of the other questionnaire.

Equipercentile linkage was used to map between different measures of mental health. The results show a stable and approximately linear mapping between RDS-4 and PHQ-9 (Fig. 4E). Additionally, our results show stable mapping between RDS-4 and N-12 (Fig. 4F), and between N-12 and GAD-7 (Fig. 4G). These results are in line with the literature showing that the personality trait of neuroticism is closely associated with mental health [Lahey, 2009].

We calculated Cronbach’s internal consistency alpha for RDS-4, which measures the internal consistency. The Cronbach alpha for RDS-4 was 0.78, which indicates a moderate to strong internal reliability. This was similar to N-12 (Cronbach alpha = 0.83).

### Mechanical Turk study to validate RDS-4

Out of 134 subjects who completed our separate validation study, 3 subjects were removed because they failed the attention questions and a further 44 subjects were removed because they completed the surveys too fast, resulting in N=87 subjects (53 female and 34 male; mean age 66.0 ± 4.8). The results showed that RDS-4 was highly correlated with other depression scales and achieved test-retest reliability comparable to other depression scales (Table 4).

**Table 4:** Comparison of RDS-4 to other depression scales from MTurk study.

| | Test-retest reliability (ICC) | Correlation with RDS-4 ($\rho$) |
| --- | --- | --- |
| RDS-4 | 0.88 | - |
| CES-D | 0.91 | 0.89 |
| PHQ-9 | 0.94 | 0.91 |
| MASQ general distress | 0.87 | 0.78 |
| MASQ anhedonic depression | 0.82 | 0.67 |
| MASQ anxious arousal | 0.92 | 0.71 |

### Exploratory brain-mental health analysis

Prior to performing the CCA, the data reduction of resting state IDPs resulted in 100 components which explained 50.1% of variance. The data reduction of the structural IDPs resulted in 24 components which explained 50.6% of variance. The data reduction of the task IDPs resulted in 2 components which explained 51.2% of variance. Therefore, the total number of brain variables input into the CCA was 126 and this was tested against the 5 mental health variables. The CCA resulted in two significant canonical covariates (R1_UV_ = 0.207, p = 0.0005 & R2_UV_ = 0.174, p = 0.015). The first multivariate canonical correlation partly replicated in the independent confirmatory sample (R1_UV(confirmatory)_ = 0.125, p = 3.7*10^−9^, where the p-value was Bonferroni corrected for the maximum of 5 canonical correlations). Although the second canonical correlation also reached significance in the confirmatory sample (R2_UV(confirmatory)_ = 0.06, p_Bonferroni_ = 0.02) we did not perform post-hoc analysis for this finding due to the low canonical correlation in the replication sample. There are a number of factors that may have contributed to the replicability of the first canonical correlation. Firstly, the CCA was relatively well-powered with 50.7 subjects per input variable leading to relatively stable estimates [Helmer et al., 2021]. Secondly, the exploratory and confirmatory samples were well matched in terms of sample characteristics. Thirdly, data reduction of IDPs prior to CCA likely reduces measurement noise. Post-hoc correlations between the averaged UV subject scores and the mental health variables and IDPs also replicated well (Fig. 5 and S2).

**Figure 5.**
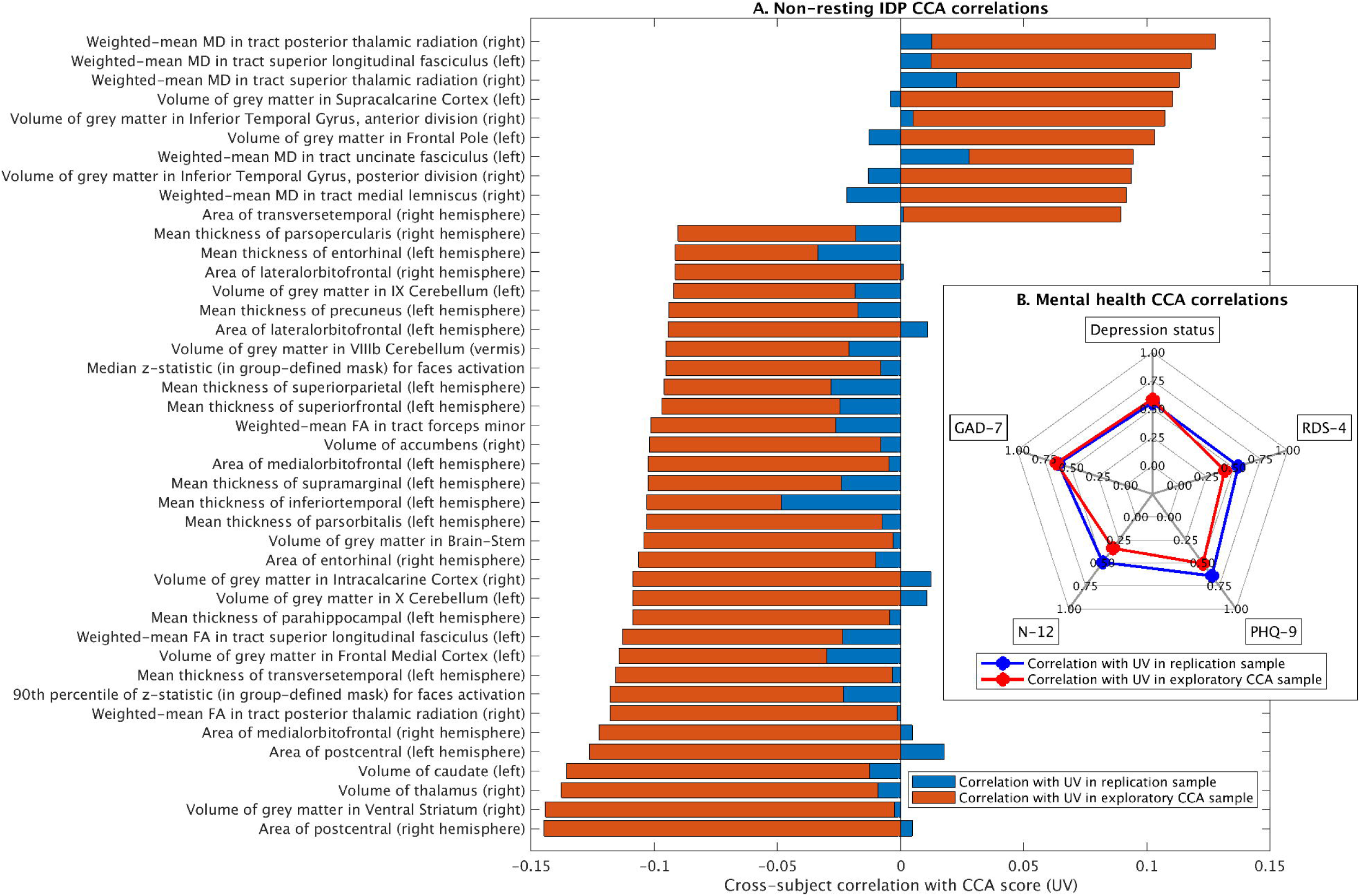
Canonical correlation results. A: post-hoc correlations for non-resting (structural & task) IDPs, showing only significant IDPs after Bonferroni correction. A similar figure for the resting state IDPs is included in the supplementary material (Fig. S2). B (insert): post-hoc CCA relations for mental health show that the first canonical covariate is broadly linked to affect-based mental health.

In terms of post-hoc correlations with IDPs, 770 resting state IDPs and 86 structural IDPs, and 1 task IDP were significantly correlated with the canonical covariate (UV) after Bonferroni correction for multiple comparisons. The post-hoc CCA results confirm many regions previously highlighted in the literature such as prefrontal and orbitofrontal cortices.

IDPs that contributed significantly to the CCA were also tested for univariate direct correlations with individual mental health variables in the independent confirmatory sample (see next section for the results). For these follow-up univariate tests, we furthermore supplemented the target IDPs with a literature-curated list (supplementary table 1) that partly overlaps with the data-driven IDP identification.

### Confirmatory analysis of effect size

Our findings showed that univariate effect sizes of the relationship between IDPs and mental health determined in our robust population sample were very low. Overall, effect sizes of the differences in the brain variables (i.e., IDPs), indicated by Cohens’ d, based on probable depression status, was larger than the Pearson’s r values from correlations between IDPs and continuous mental health measures (Fig. 6). On average, resting-state node amplitude and edge connectivity derived from partial correlation matrices appeared to have the higher effect sizes in most mental health measures, and task activity and fractional anisotropy ranked high in some mental health measures. At the level of individual IDPs, edges derived from both partial and full correlation matrices emerged as the best “predictors’’ in explaining data variance in all mental health variables except for PHQ-9 where amplitude of a few resting-state nodes ranked at top (Figs. S3-S7). These findings together suggest an overall higher effect size of resting-state in contrast to non-resting state measures on the investigated mental health variables.

**Figure 6.**
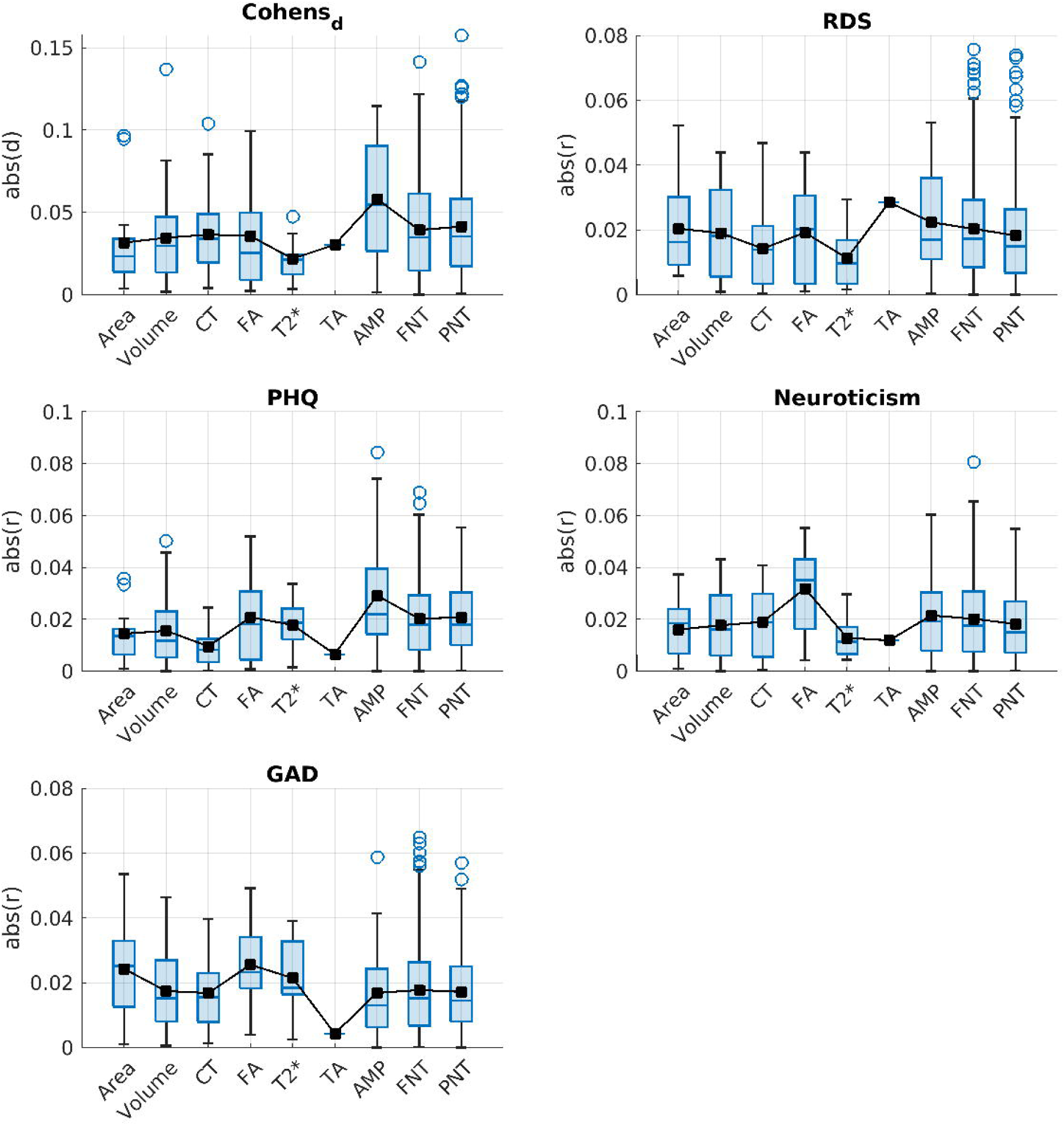
Effect sizes are shown for the grouped brain variables of structural (Area, Volume, Cortical Thickness, Fractional Anisotropy and T2*) and functional (Task Activity, Amplitude, Full Network connectivity matrix and Partial Network Connectivity Matrix) modalities. Blue boxes indicate the middle 50% of the data (i.e., the range between the first and third quartile) and small black squares and blue lines inside each box represent the mean and median values, respectively. Outliers for each grouped brain IDP are shown as blue circles, which are above the 1.5 times of inter-quartile range (IQR), indicated by the whiskers extending from the boxes. For detailed assessments of effect sizes in specific IDPs see supplementary figures S3-S7.

### Test-retest reliability of imaging measures

We next assessed the stability of IDPs over time in 624 subjects who had data from two separate scan sessions conducted approximately 2-2.5 years apart. Fig. 7A shows the distribution of inter-scan intervals for all 624 subjects. To assess test-retest reliability, ICCs were computed between the scan 1 measurements for each IDP and the corresponding scan 2 measurements for the same IDP. Then, the ICCs were assigned to categories based on the measurement modality of the corresponding IDPs: brain surface area (62 measures), brain volume (154 measures), cortical thickness (CT - 62 measures), fractional anisotropy (FA - 27 measures), mean diffusivity (MD - 27 measures), T2* value (T2 - 14 measures), task activation (TA - 16 measures), resting-state time-series amplitudes (AMP - 76 measures), full correlation-based resting-state networks (FNT - 1695 measures), and partial correlation-based resting-state networks (PNT - 1695 measures).

**Figure 7.**
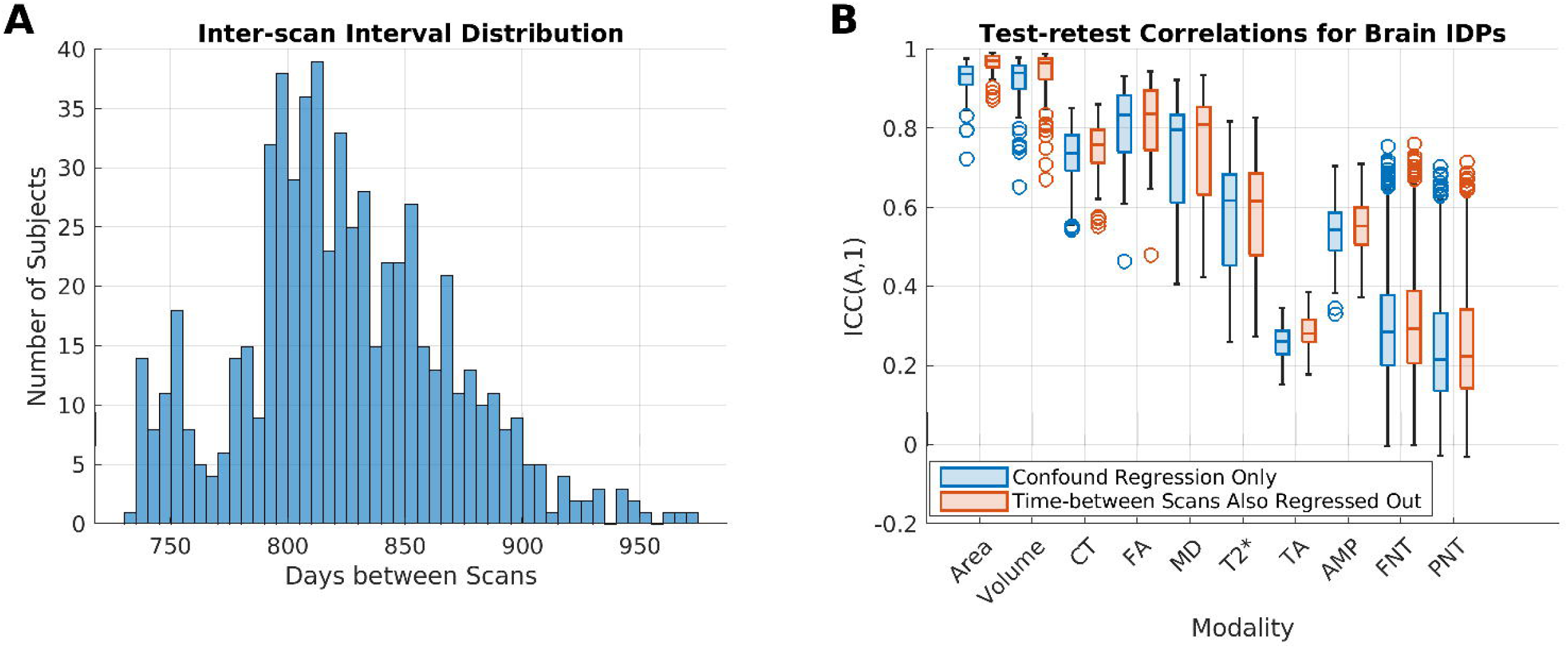
Test-retest analyses. A. The histogram shows the inter-scan interval distribution for the 624 subjects included in these analyses. The x-axis shows days between scans, and the y-axis shows the number of subjects. B. The boxplots show the ICCs obtained using brain IDPs after standard confound regression (blue) vs. ICCs obtained using brain IDPs after standard confound regression plus regressing out effects of inter-scan interval length (orange). IDP measurement modality categories are organized along the x-axis, and the y-axis shows ICC values. See also Supplemental Figure S8.

Fig. 7B depicts the distributions of ICCs for each IDP measurement modality obtained using the confound-regressed data from both scan time-points, along with those obtained after additionally regressing out the effects of inter-scan interval length (i.e. days between scans). Notably, ICC distributions were highly similar for both analyses. In general, IDPs corresponding to measures of brain structure had higher ICCs than IDPs corresponding to measures of brain function. The highest ICCs were observed for IDPs corresponding to brain volume/brain area measures and the lowest ICCs were observed for IDPs corresponding to task measures. This pattern of results is not particularly surprising since macro-scale structural properties like regional volume are expected to be relatively stable over time, especially when considering relative between-subject correlations. Macro-scale functional properties like task activation magnitudes or network connectivity patterns exhibit higher variability over time due to influences of factors such as the level of task engagement (during task), cognitive state (during rest), and physiological state (e.g. hungry vs. sated, sleepy vs. alert), and therefore are expected to have somewhat reduced test-retest stability.

Analyses performed for sub-groups of patients that did (*n*=288) vs. did not (*n*=336) exhibit changes in mental health between time-points as determined by the difference between RDS-4 measures obtained at each time point yielded highly similar results, as did those obtained after regressing out the change in RDS-4 score (See Supplementary Material section 5 and Fig. S8). Overall, these results suggest that the test-retest reliability of the IDPs is largely independent of mental health change as indicated by the RDS-4.

## 4. Discussion

In the present study we aimed to tabulate mental health questionnaires available in the UK Biobank and investigate their neural correlates. We summarize five different UKB measures of mental health: PHQ-9, GAD-7, RDS-4, N-12, and probable depression status. Our results show that all measures were moderately correlated with one another (Fig. 3). CCA analyses to identify multivariate associations between these mental health measures and IDPs indicated a significant CCA mode of covariation which linked brain IDPs to mental health scores (Fig. 5). The multivariate CCA analysis indicated a significant correlation between mental health and imaging that was largely reproducible in the independent confirmatory sample. All mental health measures contributed to the CCA result indicating a ‘trait-like’ multivariate brain-mental health association. In a separate test of univariate effect sizes, modalities with the strongest modality-mean effect sizes included amplitude and edge connectivity of resting-state networks, but univariate effect sizes were generally very low (Fig. 6). All IDPs showed moderate to high test-retest reliability, with IDPs of brain structure showing higher reliability than IDPs of brain function (Fig. 7). Together, these findings provide the foundation for future biomarkers research into mental health using the UK Biobank.

We highlighted a difference in acquisition timing of mental health questionnaires in the UKB study relative to neuroimaging data acquisition. Two well-validated measures of mental health (GAD-7 and PHQ-9) were obtained as part of the online questionnaire, which is acquired independently of scan days such that they were obtained 742 days apart (median across exploratory subjects) from scan 1 (range -1,185 to +964 days). Because of this time discrepancy (which is highly inconsistent across subjects), the PHQ-9 (which tests recent depressive symptoms over a 2-week period) is not well-suited as a state depression measure for UKB neuroimaging research despite its validity for lifetime depression [Cannon et al., 2007], and its sensitivity to depression in older populations [Levis et al., 2019]. Therefore, we introduced the RDS-4 (obtained on each day of scanning) as a new UKB measure of recently experienced depressive symptoms. We propose the RDS-4 as a more appropriate measure for any UKB neuroimaging research that aims to study acute (state) depression severity or track symptom fluctuations over time. Our results from the independent Mechanical Turk study show that the correlation between the RDS-4 and the PHQ-9 is high when obtained concurrently (0.9, Table 4), whereas a lower ‘trait-level’ correlation between RDS-4 and PHQ-9 is observed in the UKB data (0.6; Fig. 3A) due to the gap in acquisition times (Fig. S1). Furthermore, RDS-4 has high internal consistency and its scores map closely onto established measures of depression (Fig. 4, and Table 4) - further confirming its validity. The RDS-4 questions cover four different depression domains (mood, disinterest, restlessness and tiredness) that are also considered in other measures such as the Hamilton and Montgomery–Åsberg scales [Hamilton, 1967; Montgomery and Asberg, 1979]. Hence, by asking questions in different domains, the RDS-4 inventory reflects overall depression severity relatively well, despite the comparatively small number of items. The Neuroticism-12 index – also obtained on each day of scanning - is a personality trait [Eysenck and Eysenck, 1975] that is strongly related to an increased risk in depression [Hirschfeld et al., 1983; Shaw and Hare, 1969]. N-12 items assess generic traits as opposed to recently experienced clinical symptoms (RDS-4 and PHQ-9). Our results confirm that N-12 is more stable over time compared with RDS-4 and PHQ-9 as assessed by the 2-year correlation. We therefore suggest that N-12 can be used as a measure of trait-level susceptibility to depression in UKB neuroimaging research.

In terms of neuroimaging correlates of mental health, our findings show that multivariate associations explain more variance in mental health effects than univariate associations, which is supported by previous work [Marek et al., 2020]. It should be noted that our estimated effect sizes are derived from a large sample (N>2000) and are therefore expected to capture true effect sizes that are uninfluenced by sampling variability [Marek et al., 2020]. The literature to date is dominated by underpowered studies which, by design, only report high effect sizes because the significance threshold is itself high due to limited power. We have to adjust our expectations to value realistic effect sizes from well-powered samples, which may be lower but, importantly, reproducible. The observed increase in explained variance when using multivariate methods is consistent with the proposal of complex macroscopic patterns of psychopathology in mental health patients [Williams, 2016; Wise et al., 2017a]. Future biomarker research will therefore need to focus on multivariate techniques such as canonical correlation analysis, connectome fingerprinting [Finn et al., 2015], topological network properties [Zhu et al., 2017], or machine learning [Dinga et al., 2018].

One reason why multivariate methods may have higher effect sizes than univariate methods could be due to the relatively low signal-to-noise ratio and high measurement noise of individual univariate IDPs and the effective averaging that occurs in multivariate combinations of IDPs and during the dimensionality reduction prior to CCA, which reduces noise. For example, previous work showed substantial increases in heritability when combining connectivity IDPs with independent component analysis compared with univariate IDPs [Elliott et al., 2018]. Given the low SNR of individual IDPs and the risk of overfitting in multivariate methods, robust cross-validation [Poldrack et al., 2020] and independent replication of findings (in a split-half group and/or in a fully independently acquired dataset) are essential requirements for future biomarker research [Dinga et al., 2019; Dinga et al., 2020].

A second potential reason for limited effect sizes (even with the use of multivariate methods like CCA) is between-subject heterogeneity. One type of heterogeneity is diversity in symptoms, such that two patients with depression may present with largely non-overlapping symptom profiles [Drysdale et al., 2017; Feczko et al., 2019; Feczko and Fair, 2020; Kaczkurkin et al., 2020]. Another type of heterogeneity is diversity in psychophysiological disease mechanisms. Here, it is possible that the same symptom may be caused by a number of different patterns of brain changes [Feczko and Fair, 2020], which we refer to as ‘many-to-one mechanistic mapping’. Notably, both types of heterogeneity are potentially more prominent in large-scale population studies such as the UK Biobank compared with smaller studies. This is because studies with smaller samples often implement stricter exclusion criteria in relation to comorbidities and medication to control for known sources of heterogeneity. Reducing the exclusion criteria in the UKB is likely advantageous for mental health research because the UKB and other large-scale studies provide a more accurate representation of ‘real-life’ mental health as it occurs across the population. This makes the findings more likely to be generalizable.

However, gaining a better understanding of both symptom heterogeneity and many-to-one mechanistic heterogeneity is critically important for effective clinical translation of mental health biomarkers. Computational methods are available to account for heterogeneity, such as subtyping analyses to reveal any distinct sub-groups [Drysdale et al., 2017; Kaczkurkin et al., 2020] and normative modelling analysis to compare each individual against the normative range [Marquand et al., 2016]. These models of heterogeneity benefit from the large sample size available in the UK Biobank which enables stringent cross-validation.

In summary, this paper provides a guide for future neuroimaging biomarker research into affect-based mental health in the UK Biobank. We recommend using RDS-4 for imaging-based research into state depression (i.e., currently experienced symptoms) and N-12 for imaging-based research into personality traits associated with depression [Lahey, 2009]. Our results regarding the brain correlates of mental health show low effect sizes of individual IDPs, but higher effect-sizes and replicability of multivariate associations and relatively high test-retest reliability. Therefore, we recommend the use of approaches that capture multivariate patterns and parse patient heterogeneity in combination with stringent out-of-sample replication to avoid overfitting.

## Data Availability

All analysis code for this article is available at: https://github.com/PersonomicsLab/MH_in_UKB. UK Biobank data [Miller et al., 2016; Sudlow et al., 2015] are available following an access application process, for more information please see: https://www.ukbiobank.ac.uk/enable-your-research/apply-for-access. In accordance with the UKB regulations, newly derived variables in this article (e.g., RDS-4) will be made available to other researchers via UKB data access post-publication.

https://github.com/PersonomicsLab/MH_in_UKB

## Acknowledgements

We are grateful to UK Biobank and the UK Biobank participants for making the resource data possible, and to the data processing team at Oxford University for producing the shared processed data. This research was performed under UK Biobank application number 47267. This research was supported by the NIH (1 R34 NS118618-01) and the McDonnell Center for Systems Neuroscience.

## Notes

### Competing Interest Statement

The authors have declared no competing interest.

### Funding Statement

We report no external funding was received.

### Author Declarations

The Mechanical Turk section of this study was reviewed by the Washington University in St Louis IRB board and approved as exempt (IRB #201909165) because participants were fully anonymous (the option of anonymized worker IDs in CloudResearch was adopted) and no participant key was available to any member of the research team. This study also uses fully anonymized data collected previously as part of the UK Biobank and therefore does not require specific IRB approval. The UK Biobank data were acquired independently from this study with appropriate IRB approval.

### Summary of Updates

We are providing an updated manuscript revised with the changes as per reviewer comments.

