## Supplement for "Mental health in the UK Biobank: a roadmap to self-report measures and neuroimaging correlates"

### Supplementary section S1: Correlations between RDS-4 and PHQ-9 as a function of time lapsed

To investigate the effects of measurement latency on mental health measure correlation, Spearman rank correlations between the RDS-4 and PHQ-9 (both measures of depression) were computed as a function of elapsed time between measurement. Subjects were binned into subsets based on the time elapsed between the completion of assessment center information and online questionnaires to enable statistical comparisons between correlations as a function of time. Short- and long-term cutoffs were chosen to bin survey data into three time-frames. Since the questions measure experiences in the last two weeks, subjects who completed the RDS-4 and PHQ-9 within 7 days of one another constituted the “short” time-frame. The one-week period of the “short-term” bin guarantees that participants are asked to consider at least half of the same two-week period in both the PHQ-9 and the RDS-4 questionnaires. The 3-month ‘medium-term’ cut-off was determined based on the median duration of a depressive episode [[Spijker et al., 2002]](https://paperpile.com/c/OangmQ/buUwp). Hence, the short-term time-frame includes subjects whose surveys were completed between 0 and 7 days apart; the medium-term time-frame includes surveys between 8 and 92 days apart; and the long-term time-frame is composed of subjects whose surveys were taken 93 or more days apart. The significance of differences between pairs of correlations from different time-frames was computed using the Fisher transformation:

$$p=\frac{1}{2}[1+ erf(z)]$$

$z=\frac{{tanh}^{-1}(\rho_{1})-{tanh}^{-1}(\rho_{2})}{\sqrt{\frac{2}{n_{1}-3}+\frac{2}{n_{2}-3}}}$,

where $erf(z)$ is the standard error function.

The elapsed time differed widely across subjects, ranging from same-day assessment to a maximum of 1,185 days. Figure S1 shows a substantial decrease in RDS-PHQ correlation as the temporal gap between the acquisition dates widens. RDS-PHQ correlations in the “short-term” bin (one week or less; *⍴* = 0.84 ± 0.12) were significantly higher $(p=0.0002)$ than in the “long-term” bin (3 months - 3 years; *⍴* = 0.54 ± 0.01). The RDS-PHQ correlation in the medium-term gap subjects (one week - 3 months; *⍴* = 0.63 ± 0.04) also showed significant differences from the short-term $(p=0.0037)$ and long-term $(p=0.0052)$ bins.

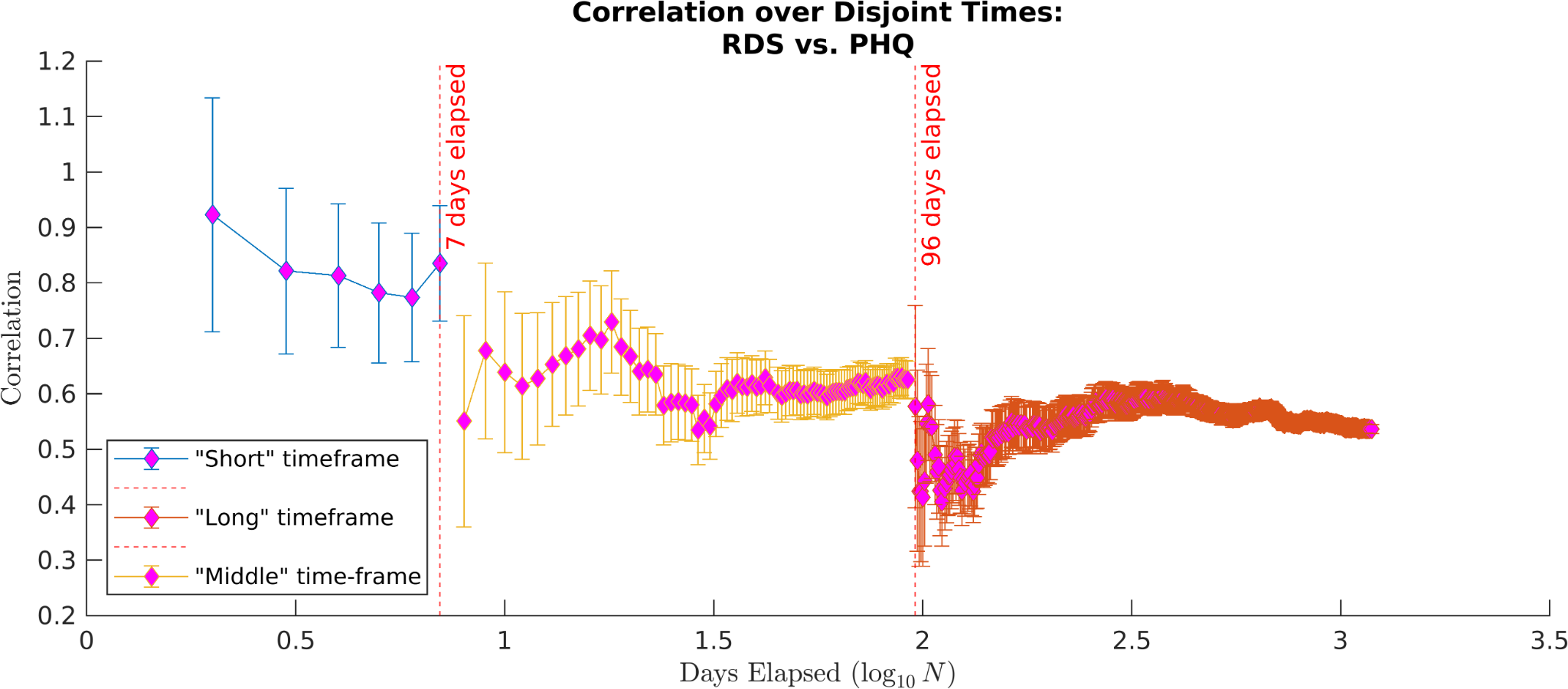

Supplementary Figure 1*.* Correlation between RDS-4 and PHQ-9 mental health variables as a function of elapsed time between measures. Correlations are binned into “short” ($\leq$7 days), “medium”, and “long” ($\geq$3 months) periods of elapsed time between surveys. For visualization purposes we computed the correlation between RDS-4 and PHQ-9 stepwise within each time-frame starting at the shortest cutoff (i.e., 0 days for the short-term time-frame, 8 days for the medium-term time-frame, and 93 days for the long-term time-frame), and incrementally relaxing the cutoff to eventually include all scores from all subjects who were measured within that time-frame. Error bars reflect the Spearman Correlation standard error at each point given the number of subjects included in the correlation.

### Supplementary section S2: literature IDP targets

|  | UKB variable ID  Left | UKB variable ID Right |
| --- | --- | --- |
| Cortical thickness in lateral PFC  Cortical thickness/volume in medial PFC  Cortical thickness/volume in OFC  Cortical thickness/volume in ACC  Cortical thickness/volume in PCC  Cortical thickness in superior parietal  Cortical thickness in inferior parietal  Volume in superior temporal gyrus  Volume in superior temporal gyrus  Volume in middle temporal gyrus  Volume in middle temporal gyrus  Volume in middle temporal gyrus  Cortical thickness in fusiform gyrus  Cortical thickness in entorhinal cortex  Cortical thickness in parahippocampal gyrus  Cortical thickness/volume in insula  Cortical volume in hippocampus  Cortical volume in amygdala  Cortical volume in thalamus  Cortical volume in striatum  White matter hyperintensity volume  FA in uncinate fasciculus  FA in superior longitudinal fasciculus  FA in forceps minor  FA in anterior thalamic radiation | 27199  25830  25846  25838  25840  27200  27179  25798  25800  25802  25804  25806  27178  27177  27187  25784  25886  25888  25878  25890  25781 (bilateral)  25513  25509  25499 (bilateral)  25490 | 27292  25831  25847  25839  25841  27293  27272  25799  25801  25803  25805  25807  27271  27270  27280  25785  25887  25889  25879  25891  -  25514  25510  -  25491 |

Supplementary Table 1: Literature-based IDP targets that were added to data-driven CCA targets, for univariate effect size tests in the confirmatory sample.

### Supplementary section S3: resting state CCA results

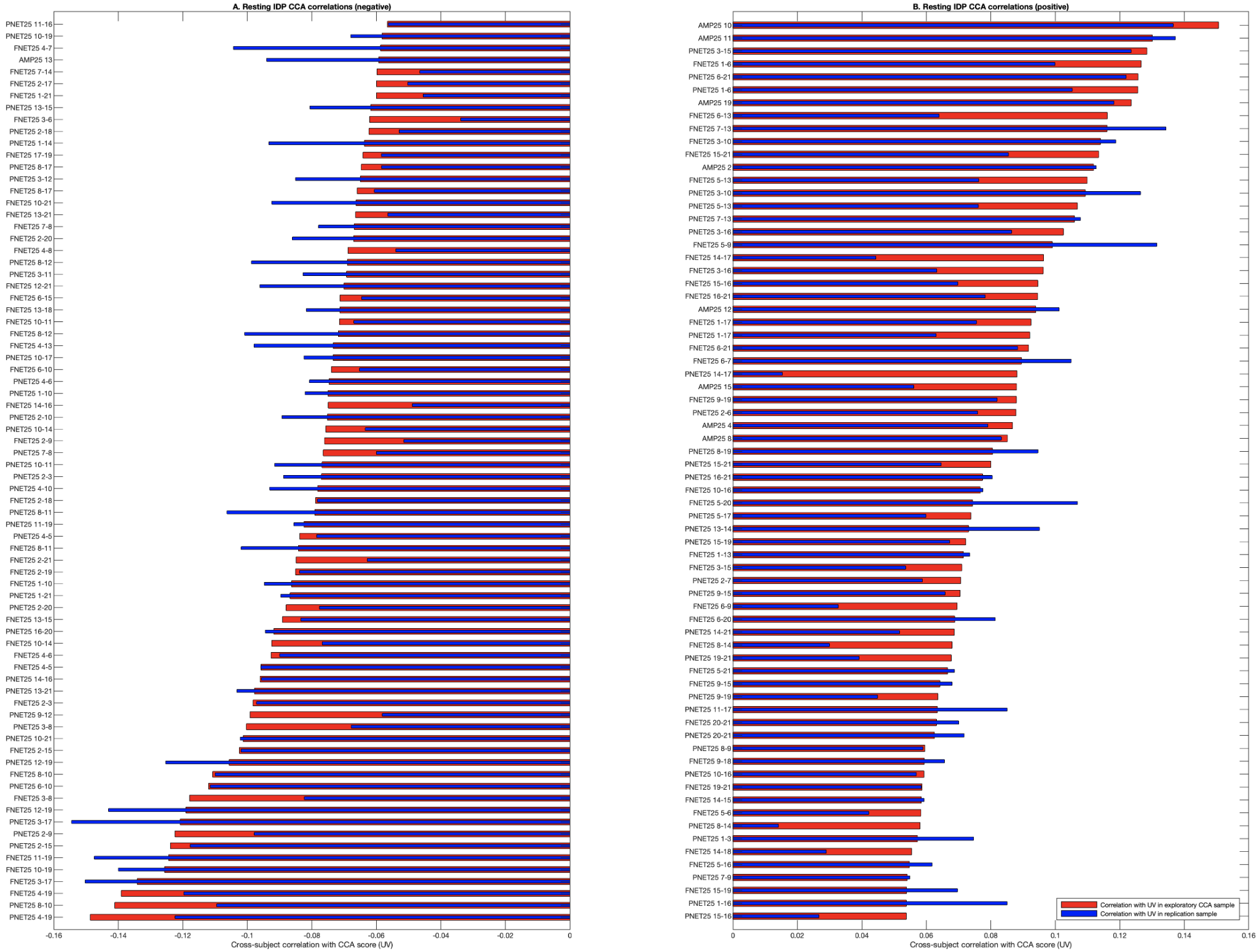

Supplementary Figure 2: Posthoc correlations for resting IDP, showing only significant IDPs after Bonferroni correction. For conciseness, only resting state IDPs from the 25 dimensional ICA are shown. Edge numbers correspond to the 3D-map browser available on <https://www.fmrib.ox.ac.uk/ukbiobank/>. Please note that CCA mental health weights were negative (Fig. 5B), so negative IDP weights (A) are positively associated with mental health and positive IDP weights (B) are negatively associated with mental health.

### Supplementary section S4: Additional IDP effect size results

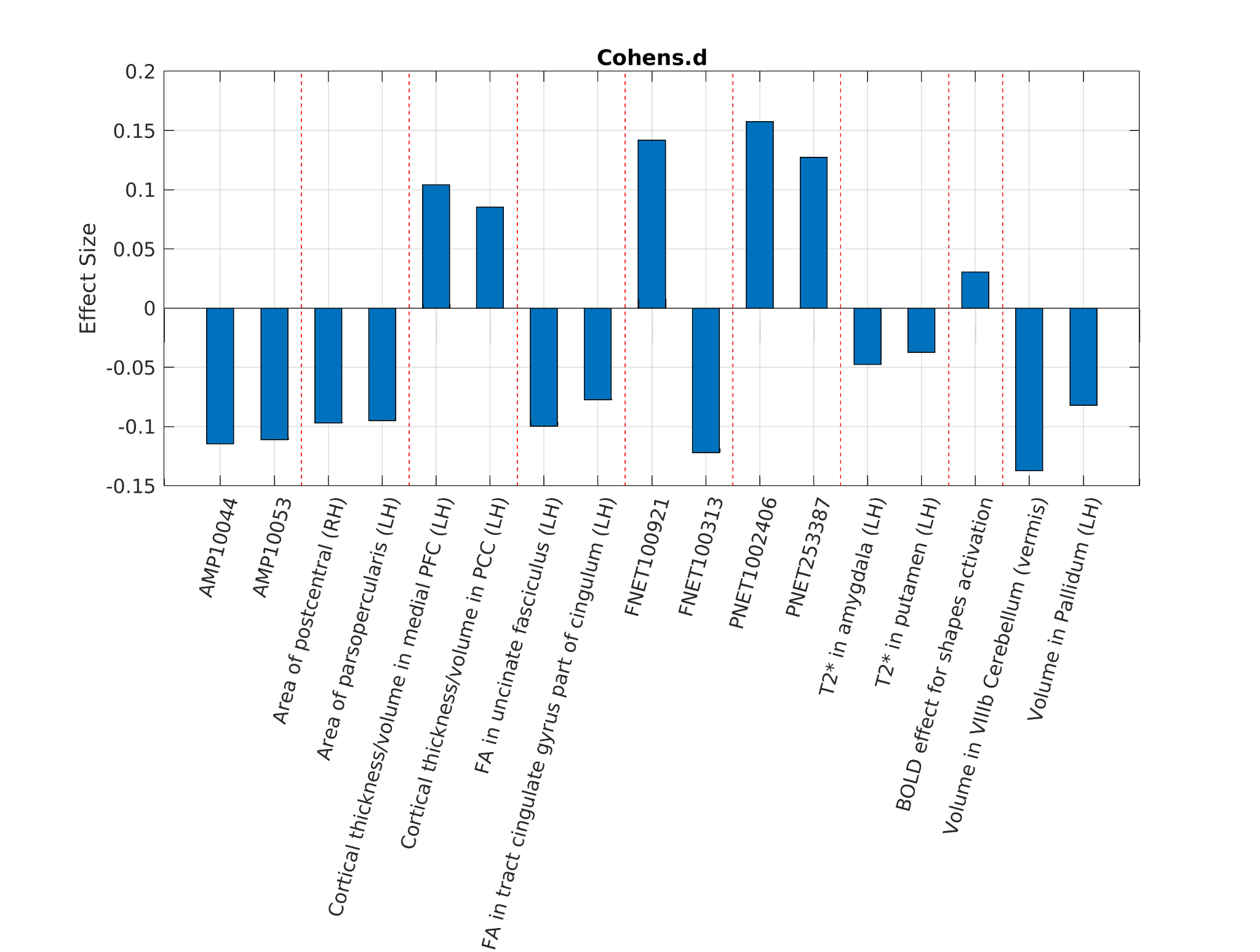

Supplementary Figure 3. Cohen’s d for top 2 brain variables (IDPs) from each individual modality based on group comparisons of probable depression status, as separated by red dash lines in the figure. Within each modality, these individual IDPs were most effective in differentiating the probable depression status and not probable depression status participants. Grouped modalities from left to right are in order of Amplitude, Area, Cortical Thickness, Fractional Anisotropy, Full Correlation Matrix, Partial Correlation Matrix, T2*, Task Activity and Volume. Note that the task activity has only one IDP available (i.e., BOLD effect for shapes activity).

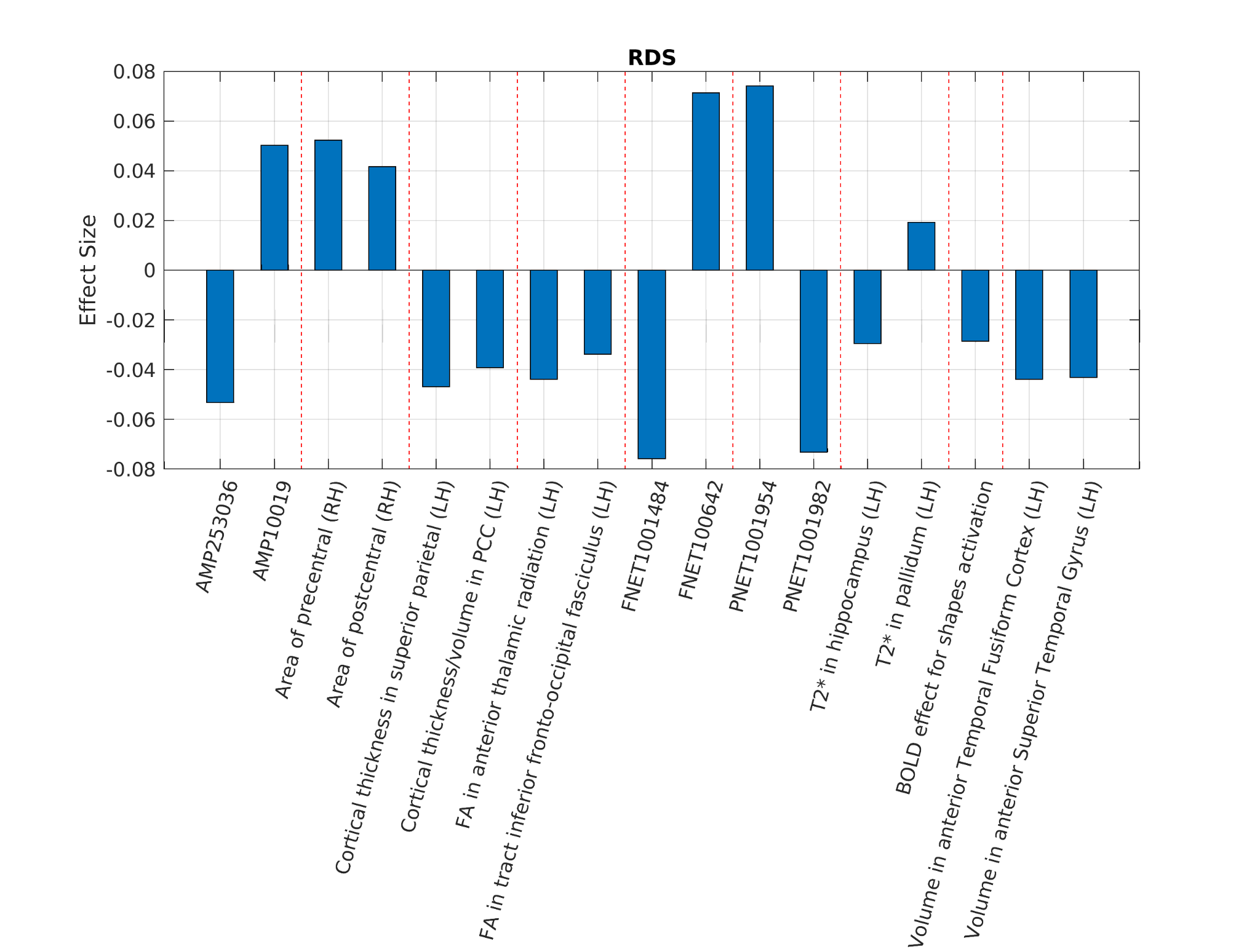

Supplementary Figure 4. Correlation coefficients (Pearson’s r) for top 2 brain variables (IDPs) from each individual modality associated with RDS-4, as separated by red dash lines in the figure. These IDPs showed the largest effect sizes within each modality in explaining variance of RDS-4 measure. Grouped modalities from left to right are in order of Amplitude, Area, Cortical Thickness, Fractional Anisotropy, Full Correlation Matrix, Partial Correlation Matrix, T2*, Task Activity and Volume. Note that the task activity has only one IDP available (i.e., BOLD effect for shapes activity).

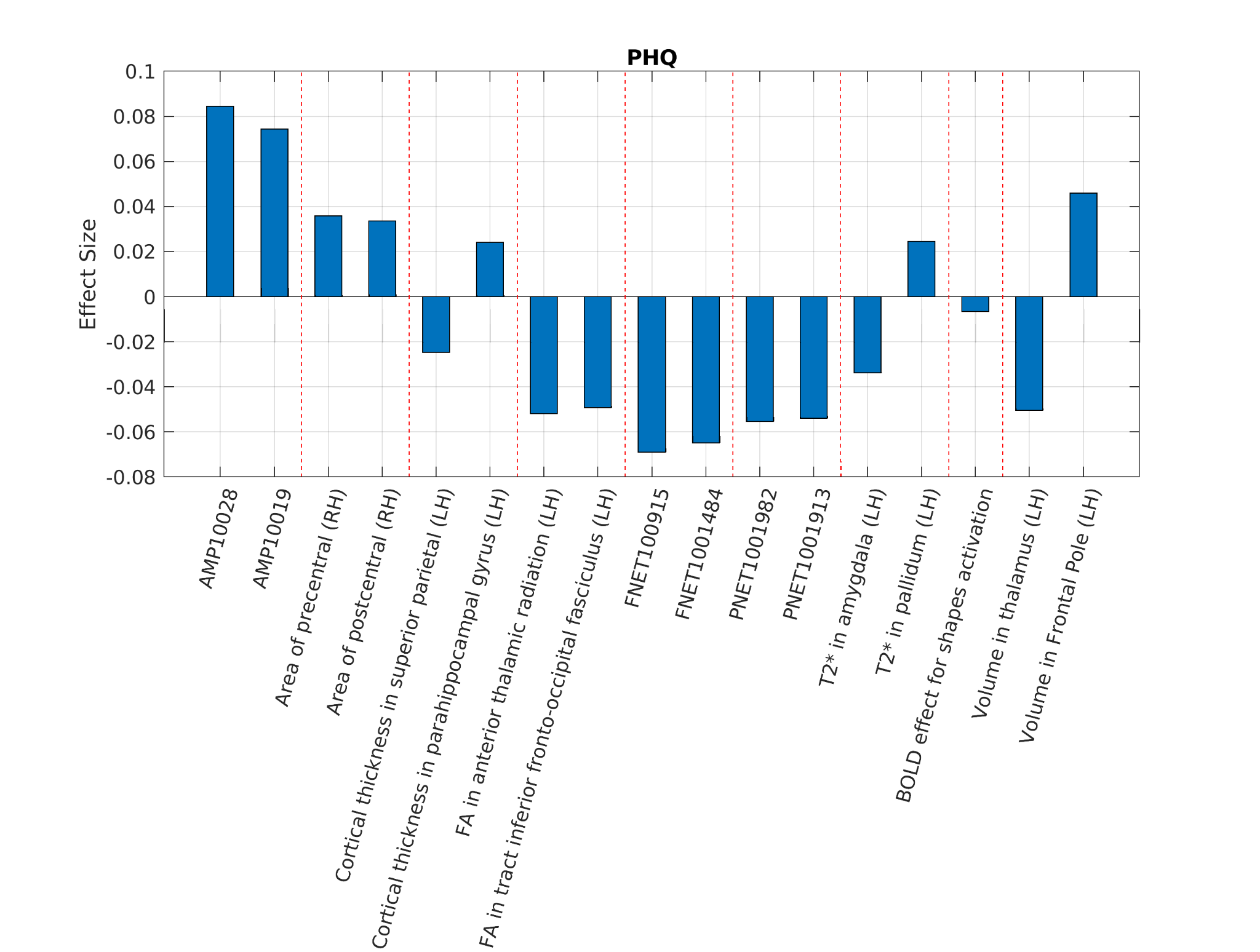

Supplementary Figure 5. Correlation coefficients (Pearson’s r) for top 2 brain variables (IDPs) from each individual modality associated with PHQ-9, as separated by red dash lines in the figure. These IDPs showed the largest effect sizes within each modality in explaining variance of PHQ-9 measure. Grouped modalities from left to right are in order of Amplitude, Area, Cortical Thickness, Fractional Anisotropy, Full Correlation Matrix, Partial Correlation Matrix, T2*, Task Activity and Volume. Note that the task activity has only one IDP available (i.e., BOLD effect for shapes activity).

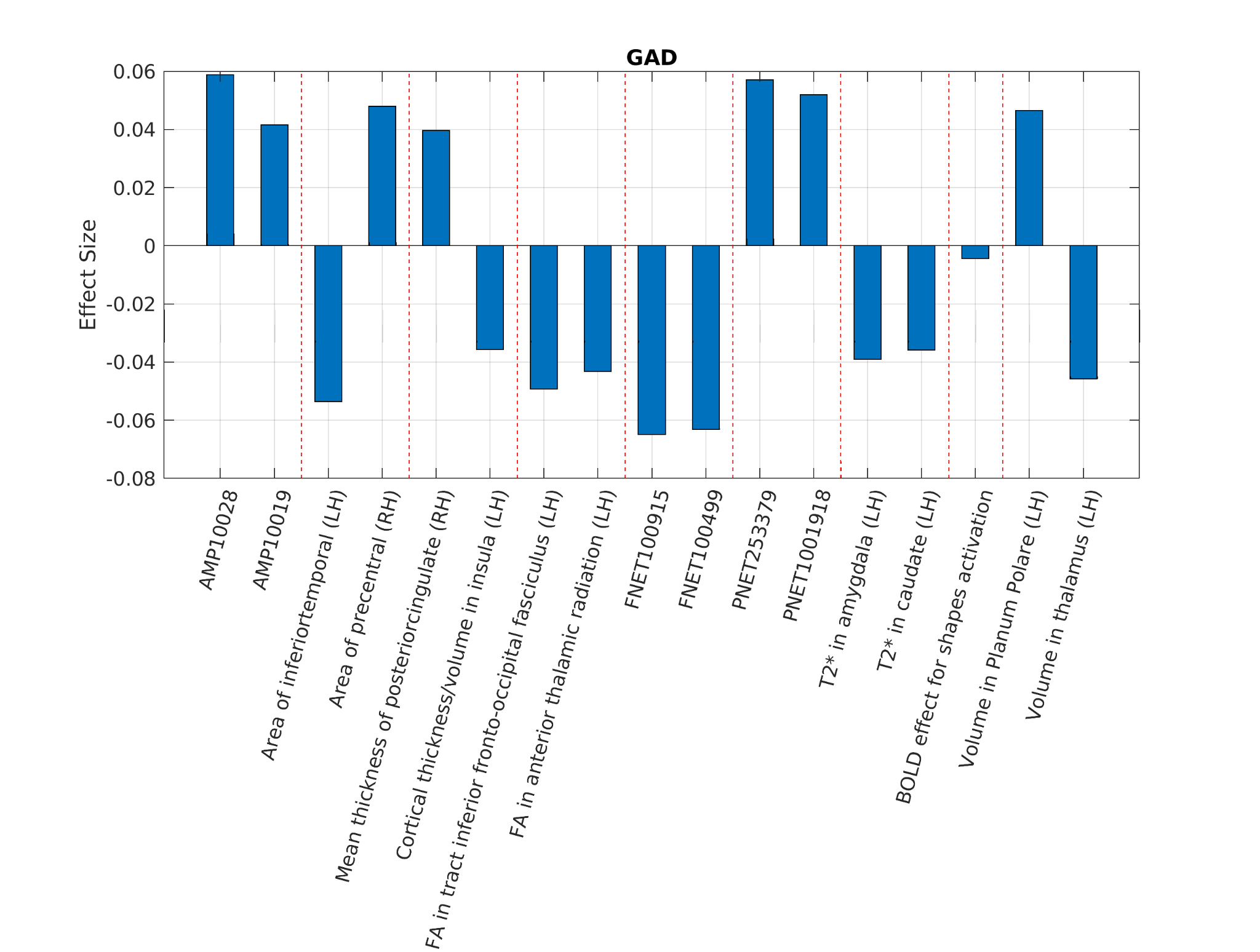

Supplementary Figure 6. Correlation coefficients (Pearson’s r) for top 2 brain variables (IDPs) from each individual modality associated with GAD-7, as separated by red dash lines in the figure. These IDPs showed the largest effect sizes within each modality in explaining variance of GAD-7 measure. Grouped modalities from left to right are in order of Amplitude, Area, Cortical Thickness, Fractional Anisotropy, Full Correlation Matrix, Partial Correlation Matrix, T2*, Task Activity and Volume. Note that the task activity has only one IDP available (i.e., BOLD effect for shapes activity).

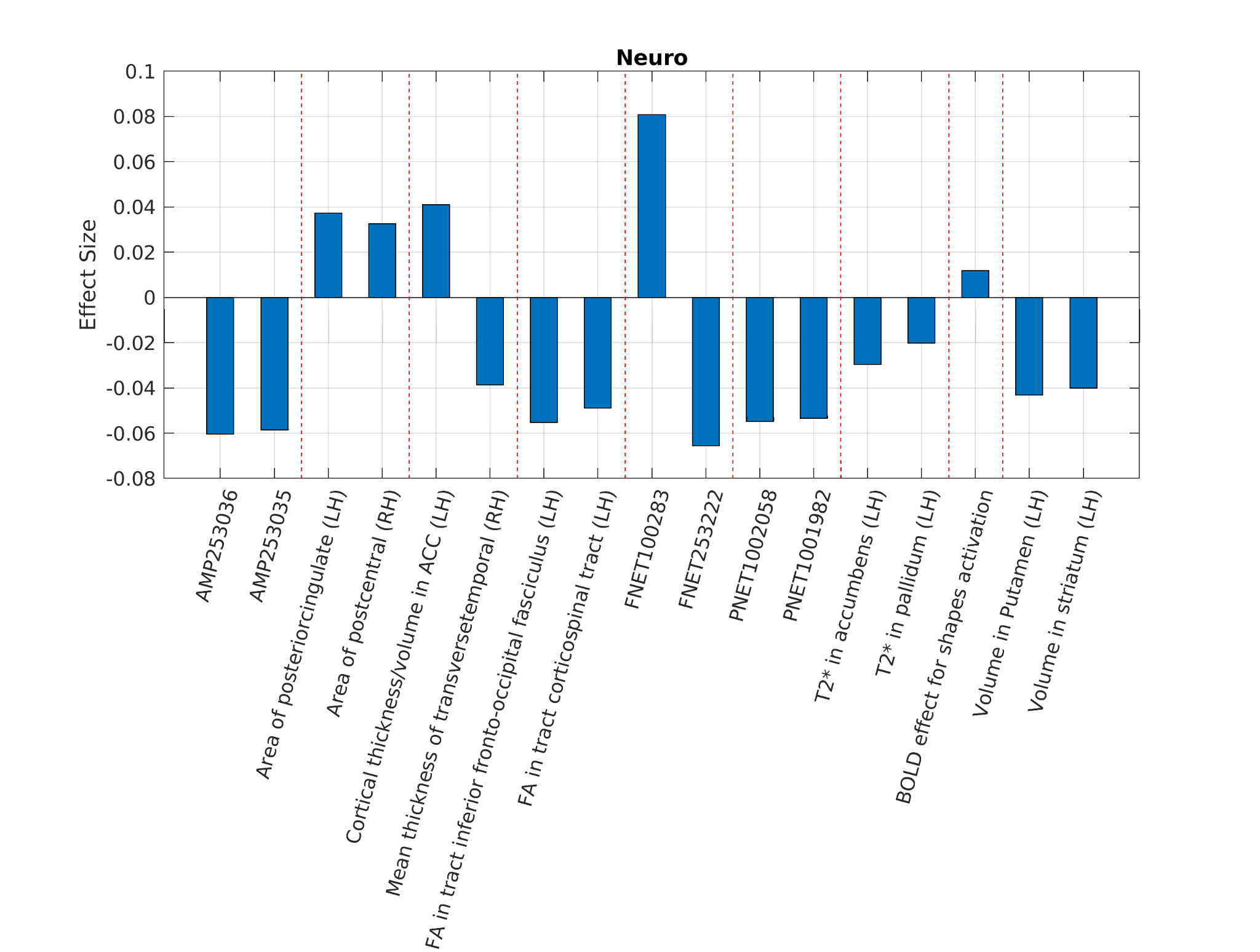

Supplementary Figure 7. Correlation coefficients (Pearson’s r) for top 2 brain variables (IDPs) from each individual modality associated with Neuroticism (N-12), as separated by red dash lines in the figure. These IDPs showed the largest effect sizes within each modality in explaining variance of Neuroticism-12 measure. Grouped modalities from left to right are in order of Amplitude, Area, Cortical Thickness, Fractional Anisotropy, Full Correlation Matrix, Partial Correlation Matrix, T2*, Task Activity and Volume. Note that the task activity has only one IDP available (i.e., BOLD effect for shapes activity).

### Supplementary section S5: Additional test-retest reliability results

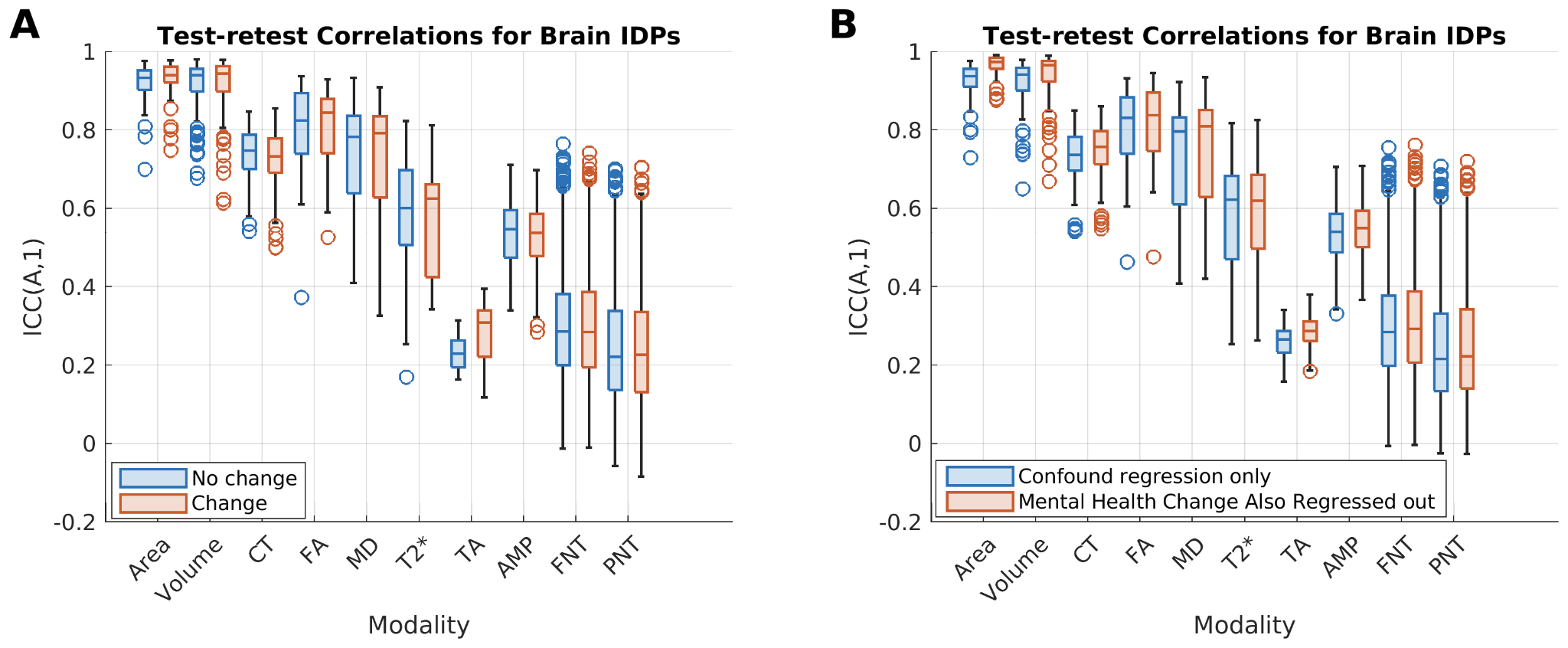

Supplementary Figure 8. Test-retest correlations for brain IDPs was not influenced by mental health. A) Boxplots show test-retest correlations as in Figure 7B in the main manuscript, but box colors correspond to test-retest correlation distributions obtained from separate analyses of the No Change (blue) and Change (orange) sub-groups. Subjects with identical RDS-4 scores between time point 1 and time point 2 were in the No Change subgroup, and subjects with one-or-more point difference were in the Change subgroup. B) Regressing out the change in RDS-4 score from IDP data for timepoint 1 and for timepoint 2 resulted in a very minor increase in the test-retest reliability across all IDP categories.
